# Isolating the effect of COVID-19 related disruptions on HIV diagnoses in the United States in 2020

**DOI:** 10.1101/2022.08.30.22279222

**Authors:** Alex Viguerie, Ruiguang Song, Anna Satcher Johnson, Cynthia M. Lyles, Angela Hernandez, Paul G. Farnham

**Author notes:** Disclaimer: The data used for this analysis was collected as part of the public health program activity called PS18-1802 Integrated HIV Surveillance and Prevention Programs for Health Departments (Component A), which is a routine disease surveillance activity across 60 jurisdictions throughout the U.S. A project determination made by the National Center for HIV, Hepatitis, STD, and TB Prevention (NCHHSTP) on behalf of The Centers for Disease Control and Prevention (CDC) on January 22, 2018 determined this project to be exempt from needing IRB review as it is not human subjects research. The National Center for HIV, Hepatitis, STD, and TB Prevention (NCHHSTP) on behalf of The Centers for Disease Control and Prevention (CDC) approved the use of this data in the current document on August 25, 2022.

## Abstract

**Background:** Diagnoses of HIV in the US decreased by 17% in 2020 due to COVID-related disruptions. The extent to which this decrease is attributable to changes in HIV testing versus HIV transmission is unclear. We seek to better understand this issue by analyzing the discrepancy in expected versus observed HIV diagnoses in 2020 among persons who acquired HIV between 2010-2019, as changes in diagnosis patterns in this cohort cannot be attributed to changes in transmission.

**Methods:** We developed three methods based on the CD4-depletion model to estimate excess missed diagnoses in 2020 among persons with HIV (PWH) infected from 2010-2019. We stratified the results by transmission group, sex assigned at birth, race/ethnicity, and region to examine differences by group and confirm the reliability of our estimates. We performed similar analyses projecting diagnoses in 2019 among PWH infected from 2010-2018 to evaluate the accuracy of our methods against surveillance data.

**Results:** There were approximately 3100-3300 (approximately 18%) fewer diagnoses than expected in 2020 among PWH infected from 2010-2019. Females (at birth), heterosexuals, persons who inject drugs, and Hispanic/Latino PWH missed diagnoses at higher levels than the overall population. Validation and stratification analyses confirmed the accuracy and reliability of our estimates.

**Conclusions:** The substantial drop in number of previously infected PWH diagnosed in 2020, suggests that changes in testing played a substantial role in the observed decrease. Levels of missed diagnoses differed substantially across population subgroups. Increasing testing efforts and innovative strategies to reach undiagnosed PWH are needed to offset this diagnosis gap. These analyses may be used to inform future estimates of HIV transmission during the COVID-19 pandemic.

## Introduction

The outbreak of COVID-19 in 2020 has significantly disrupted the HIV continuum of care in the United States. Recent work has shown that COVID-19-related disruptions have affected Pre-exposure prophylaxis (PrEP) uptake and adherence [1], antiretroviral therapy (ART) uptake and adherence [2], and HIV testing and care provision [3], [4]. It is therefore not surprising that HIV diagnoses in the United States decreased substantially in 2020, with official data suggesting a drop of approximately 17% from 2019 [5], [6].

However, it is unclear to what extent the drop in diagnoses is attributable to reductions in persons seeking testing versus reductions in transmission (incidence), resulting in fewer persons requiring a diagnosis. Both effects have likely contributed; however, estimating their relative magnitudes is not straightforward. This lack of clarity has caused problems in HIV surveillance, as most models used to estimate incidence are derived from diagnosis data [7]– [9]. Uncertainty regarding the interpretation of 2020 diagnosis data has called many of the underlying assumptions in such models into question. For this reason, the Centers for Disease Control and Prevention (CDC) has not published official incidence estimates for 2020. While the diagnosis data have been published, CDC has urged caution in the interpretation of these data [5], [6].

In the absence of an incidence estimate, modeling studies have attempted to quantify both present and future effects of COVID-19 on trends in HIV by running multiple simulations under varying assumptions on changes in transmission behaviors and care provision [10]–[12]. While such studies have value, the lack of clarity around these relative attribution issues limits their utility, as the projected long-term effects may vary substantially depending on which assumptions are used. It is therefore critical to better understand the proximate causes of this drop in diagnoses.

In the last decade, CD4-measurements taken at diagnosis have been widely employed to estimate year of HIV infection. Such measurements form the basis of modern incidence-estimation methods [7]–[9], [13], [14]. Unpublished data from CDC’s National HIV Surveillance System (NHSS) show that, historically, 60-70% of HIV infections are *not* diagnosed within the first calendar year of infection. Additionally, each year, only 30-35% of new HIV diagnoses result from infections within that same year. Thus, our goal is to estimate the discrepancy between expected and observed diagnoses in 2020 (referred to as *excess missed diagnoses*) among PWH infected in or before 2019. We use the terminology *excess missed diagnoses*, as each year there are already many missed diagnoses, and hence, we are referring to additional missed diagnoses above the expected level.

By restricting our cohort to persons infected before 2020, we ensure that any observed drop in diagnoses in this group *cannot* be attributed to changes in incidence, as such persons were infected *before* the beginning of the COVID-19 pandemic. By removing uncertainty around incidence, we can provide a reliable estimate of what is apparently a large portion of PWH unaware of their status due to COVID-19. Additionally, focusing on this population allows us to better quantify the effect of COVID-19-related testing disruption on HIV diagnosis in 2020.

## Methods

In this section, we provide a brief description of the three different methods we developed, and the data used for estimating excess missed HIV diagnoses in 2020. The full mathematical details of all methods are provided in Appendix A. Data from CDC’s National HIV Surveillance System were used for all analyses. Collection of these data, complete since 2009 and which are used to construct annual incidence estimates, provide the annual number of new HIV diagnoses for each infection year (as estimated by the CD4-depletion model) [8], [9], [15]–[17].

A minority, ranging from 10 – 15%, of new diagnoses do not have an associated CD4 measurement. To account for this limitation, we assumed that the infection year-distribution of new diagnoses without a CD4 measurement is identical to that with a CD4 measurement. This assumption is also employed in incidence estimation [7]–[9].

Some of the proposed methods require incidence data as an input. In such cases, we used incidence estimates published by CDC for years 2009-2019 [15]–[17].

### Diagnosis-based method

For the diagnosis-based method, we used regression analysis to project the number of HIV diagnoses in 2020 based on pre-COVID diagnosis trends [18]. The difference between the projected number and the observed number of diagnoses in 2020 is the number of excess missed diagnoses. We distributed these diagnoses by year of infection based on the distribution among the observed diagnoses in 2020.

More specifically, we used a 4-year log-normal linear regression model to project the number of HIV diagnoses in 2020 based on diagnosis trends in 2016-2019. To derive the infection-year distribution, each HIV case diagnosed in 2020 was weighted to account for those excess missed diagnoses in 2020. We calculated this weight as the projected number of diagnoses divided by the observed number of HIV diagnoses in 2020. We stratified HIV cases diagnosed in 2020 by year of HIV infection based on their first CD4 test after diagnosis. We derived the excess missed diagnoses by year of infection from the increased case counts based on weighted and unweighted numbers, where the unweighted case counts sum to the total observed number of HIV diagnoses in 2020 and the weighted case counts sum to the total projected number of HIV diagnoses in 2020

We reduced the estimated number of excess missed diagnoses with infection in 2020 by 50%, and we redistributed them proportionally to those with infection years prior to 2020. This adjustment was motivated by the fact that infections that occurred before 2020 may conceivably be diagnosed at any point during 2020. However, infections that occurred during 2020 can only be diagnosed post-infection, reducing the available time to be diagnosed by 6 months on average.

### Incidence-based method

Our second method for estimating excess missed diagnoses, the incidence-based method, incorporates incidence data in addition to diagnosis data. The incidence-based method works by projecting diagnoses in a year *y* among PWH infected in a year *x* as follows:

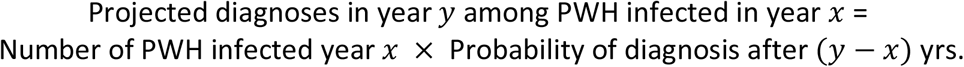

The number of excess missed diagnoses in year *y* among those infected in *x* is then:

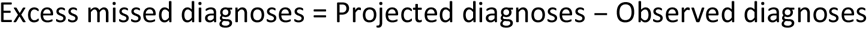

For the total number of PWH infected in year *x*, we used CDC U.S. HIV incidence estimates for year *x* from CDC’s NHSS [15]–[17].

To estimate the probability of diagnosis *y* − *x* years after infection, we used both incidence and diagnosis data. For example, define the estimate of the probability of diagnosis in 2020 of PWH infected in 2017, or 3 years after infection, as:

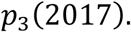

We first computed the probability of diagnosis after 3 years for PWH infected in the years preceding 2017 with NHSS data. For example:

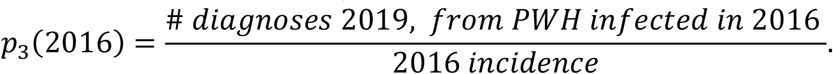

From our estimates: *p*_3_(2016), *p*_3_(2015), *p*_3_(2014), … we can fit a function 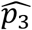, which allows us to extrapolate the expected probability of diagnosis in 2020 for PWH infected in 2017. Then:

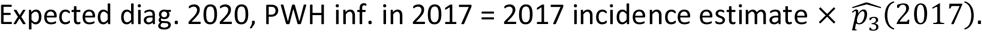

The difference between the expected and observed diagnoses among PWH infected in 2017 gave us the excess missed diagnoses in 2020 for 2017 infections. We repeated this process for infection years for which we had reliable data (beginning in 2010). Summing these results, we obtained our estimate for total excess missed diagnoses in 2020.

As with any function fitted to measured data, 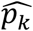 may be defined in different ways. In the current analysis, we considered two ways of defining 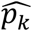: by linear regression and by averaging. For linear regression, we applied a robust least-squares linear fitting with surveillance data for 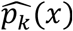, the probability of diagnosis after *k* years in a diagnosis year *x*. For the averaging approach, we defined 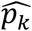 as a constant function, as the average (arithmetic mean) probability over the last 3 years of diagnosis after *k* years.

### Diagnosis-delay based method

This method is structurally similar to the incidence-based method, as it also estimates the number of diagnoses in a year *y* among PWH infected in a year *x* as follows:

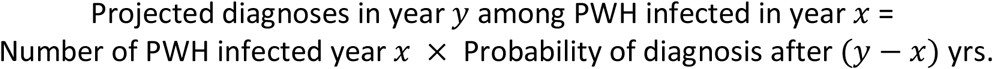

However, the diagnosis-delay based method is distinct from the incidence-based method in two key respects. First, instead of assuming known HIV incidence in years prior to 2020, we estimated the annual number of infections based on the observed CD4 data, but we limited the analysis to HIV cases diagnosed up to 2019, and we used a simplified estimation procedure (with case counts by calendar year). This method is the same as the one used to estimate HIV incidence at the national level [8], [19].

Second, the method of estimating the probability of diagnosis is different from the incidence-based method. In the diagnosis-delay based method, we estimated this probability through a conditional probability analysis. We write the cumulative probability that an infection acquired in year 0 is diagnosed by year *k* as 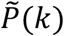. We estimated 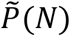 for a given year *N* (in this case *N* = 12, the longest possible period for which our data are complete) and then obtained 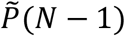 through conditional probability:

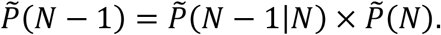

Repeating the procedure:

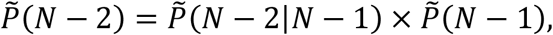

similarly for *N* − 3, *N* − 4, and so forth.

The 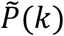 are cumulative probabilities, which give the probability of diagnosis *within k* years of infection. However, we need the *annual* diagnosis probability-the probability of diagnosis exactly *k* years after infection, 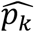. This is promptly obtained as:

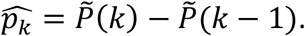

## Results

### Total Missed HIV Diagnoses in 2020

Our estimates of the total number of excess missed diagnoses in 2020 for PWH (aged 13 or older) infected from 2010-2019 ranged from **3154-3315** for our different estimation methods (Figure 1, Table 1). This is approximately a 18% decline from the expected number of ∼18,000 HIV diagnoses in 2020. These estimates were consistent across all methods, with the differences between them 5% or less. Additionally, the different methods showed similar results on infection year-distribution, with the correlation coefficients between the different methods .92 or higher.

**Table 1:** Excess missed HIV diagnoses in 2020, by infection year, and total, for each method, United States.

| *Infection year* | 2010 | 2011 | 2012 | 2013 | 2014 | 2015 | 2016 | 2017 | 2018 | 2019 | *Total* | *Total expected* |
| --- | --- | --- | --- | --- | --- | --- | --- | --- | --- | --- | --- | --- |
| <u>Diag.-based</u> | 169 | 240 | 260 | 286 | 333 | 349 | 371 | 383 | 386 | 376 | **3154** | **17850** |
| <u>Diag. delay-based</u> | 201 | 204 | 245 | 330 | 318 | 401 | 440 | 403 | 409 | 364 | **3315** | **18011** |
| <u>Inc.-based (linear fit)</u> | 188 | 200 | 238 | 334 | 331 | 411 | 428 | 377 | 396 | 335 | **3238** | **17934** |
| <u>Inc.-based (averaged)</u> | 188 | 200 | 242 | 329 | 315 | 403 | 449 | 397 | 416 | 349 | **3288** | **17984** |

**Figure 1:**
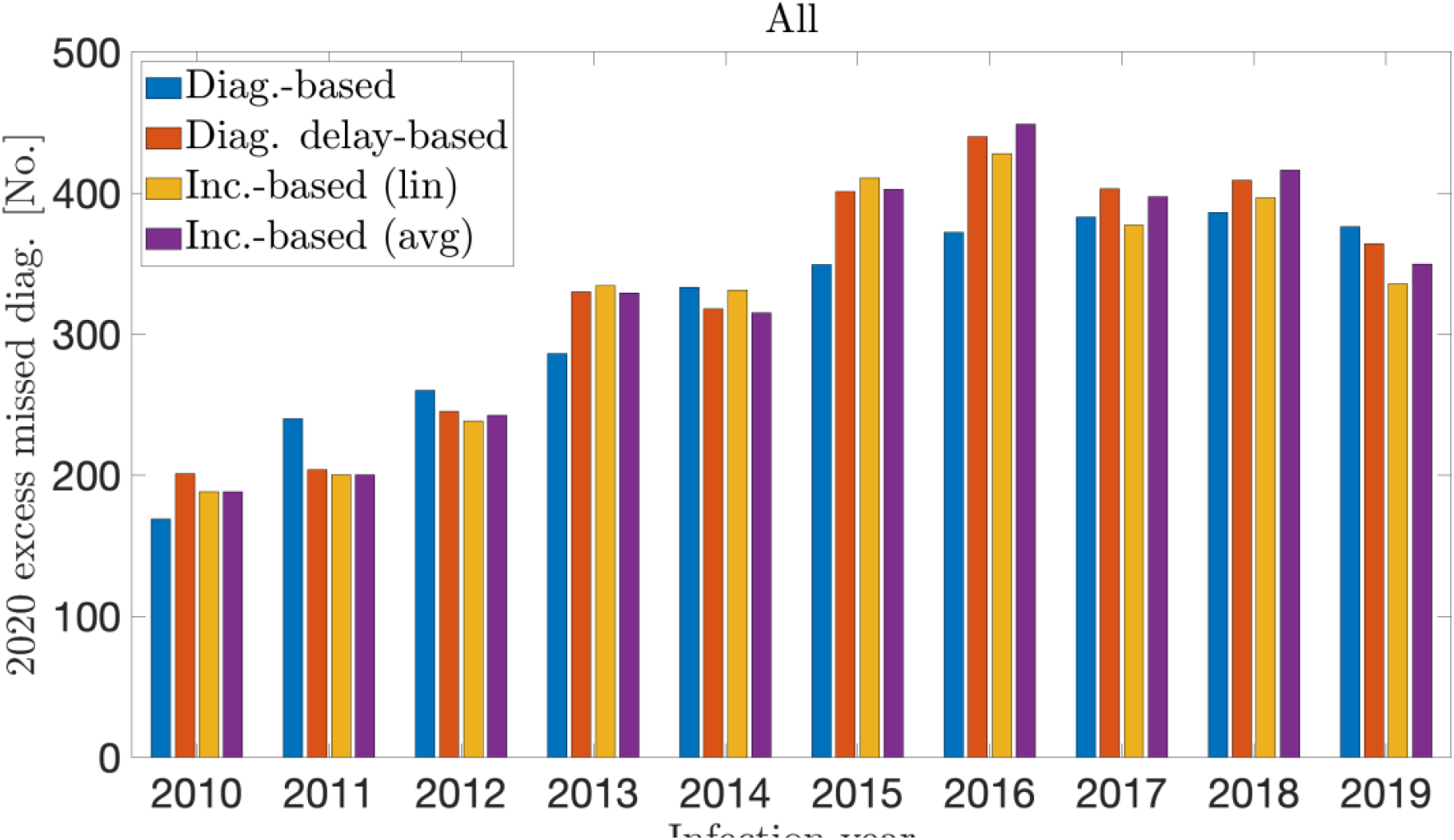
Excess missed HIV diagnoses in 2020, by infection year for each method, United States

In the diagnosis delay- and incidence-based (averaged) methods, we estimated that the highest concentration of excess missed diagnoses in 2020 occurred in the infection years 2015-2018, that is, among persons infected between 2 to 5 years prior to 2020. There was a more even infection-year distribution in the diagnosis-delay based and incidence based (linear fit) methods. However, these methods similarly projected high concentrations of excess undiagnosed infections occurring in the years 2016-2019.

### Stratification by transmission group

In Figure 2 and Table 2, we show the estimated excess missed diagnoses stratified by transmission group: men who have sex with men (MSM), heterosexuals (HET), persons who inject drugs (PWID), and men who have sex with men who also inject drugs (MWID). The results are organized by method and infection year. We estimated 1726-2034 excess missed diagnoses among MSM, 741-867 among heterosexuals, 263-446 among PWID, and 207-216 among MWID.

**Table 2:** Excess missed HIV diagnoses in 2020, among persons infected during 2010-2019, by transmission group, for each method, United States

| Transmission group | Diag.-based | Diag. delay-based | Inc.-based (linear fit) | Inc.-based (averaged) |
| --- | --- | --- | --- | --- |
| MSM | 1726 | 1995 | 1737 | 2034 |
| HET | 741 | 867 | 854 | 828 |
| PWID | 446 | 283 | 370 | 263 |
| MWID | 216 | 212 | 209 | 207 |

**Figure 2:**
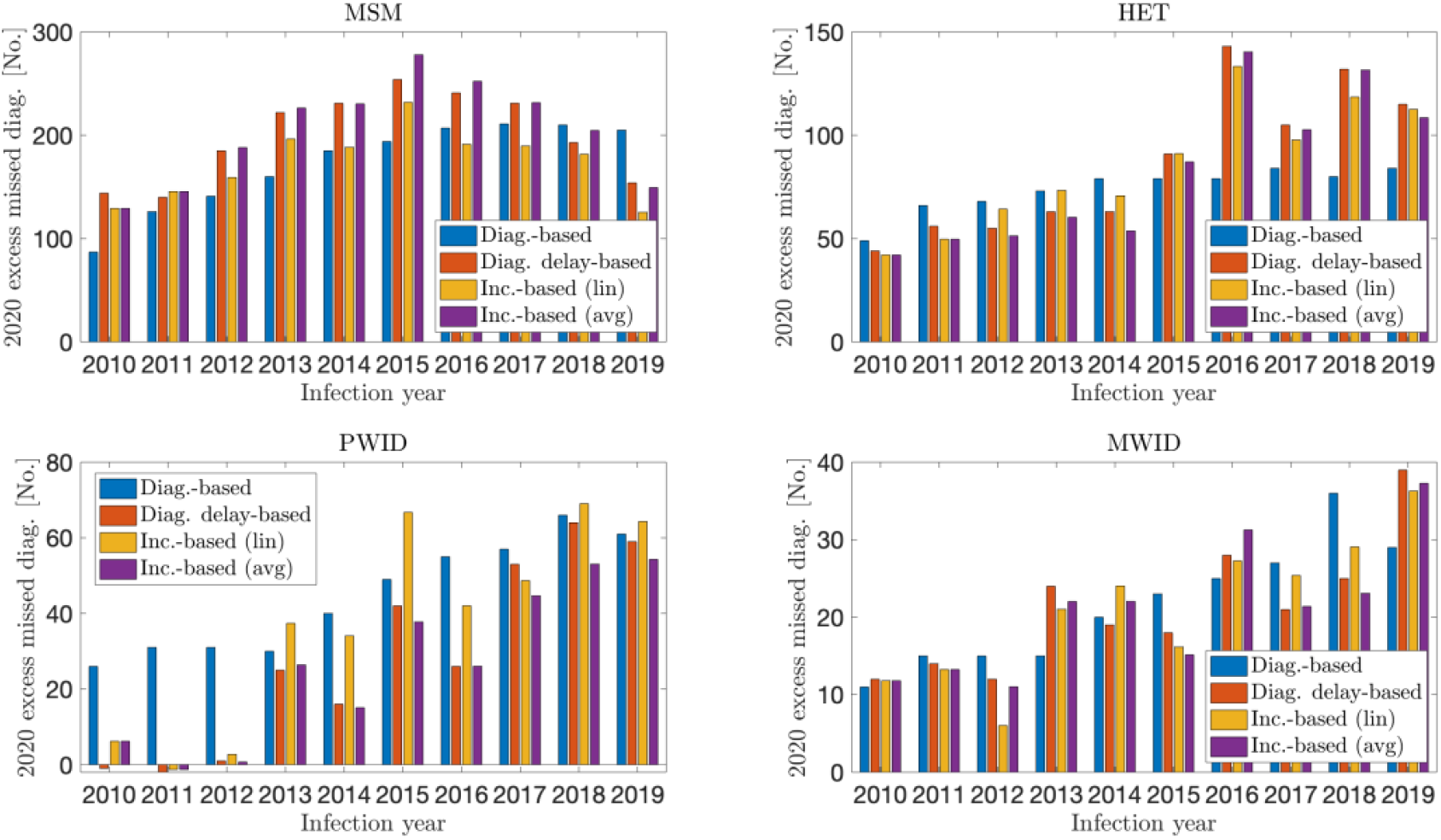
Excess missed HIV diagnoses in 2020, by infection year and transmission group, United States. Comparison between methods.

Missed diagnoses among MSM showed a regular distribution by infection year. Somewhat higher levels of missed diagnoses were observed among infections from 2014-2016, and somewhat lower levels among older and more recent infections. Among MWID, we observed an asymmetric trend, with more recent infections missing diagnoses at substantially higher levels compared to older infections (∼60% of estimated missed diagnoses from infection years 2016-2019).

The incidence and diagnosis delay-based methods showed virtually no diagnoses missed among PWID infected from 2010-2012. Among heterosexuals, the incidence and diagnosis-delay based methods showed a marked infection-year trend, with missed diagnoses occurring at a notably higher level among PWH infected more recently (∼60% of estimated missed diagnoses from infection years 2016-2019).

Figure 3 (a) displays the number of expected HIV diagnoses in 2020, broken down by observed versus missed, overall and by each transmission group (left axis), as well as the percent of expected HIV diagnoses that were missed within each transmission group (right axis). For all PWH infected from 2010-2019, 18% (∼3288) of the approximately 18,000 expected HIV diagnoses in 2020 were missed^1^. This percentage was slightly lower for MSM (16%), somewhat higher for PWID (19%), and substantially higher for heterosexuals (24%) and MWID (31%).

**Figure 3:**
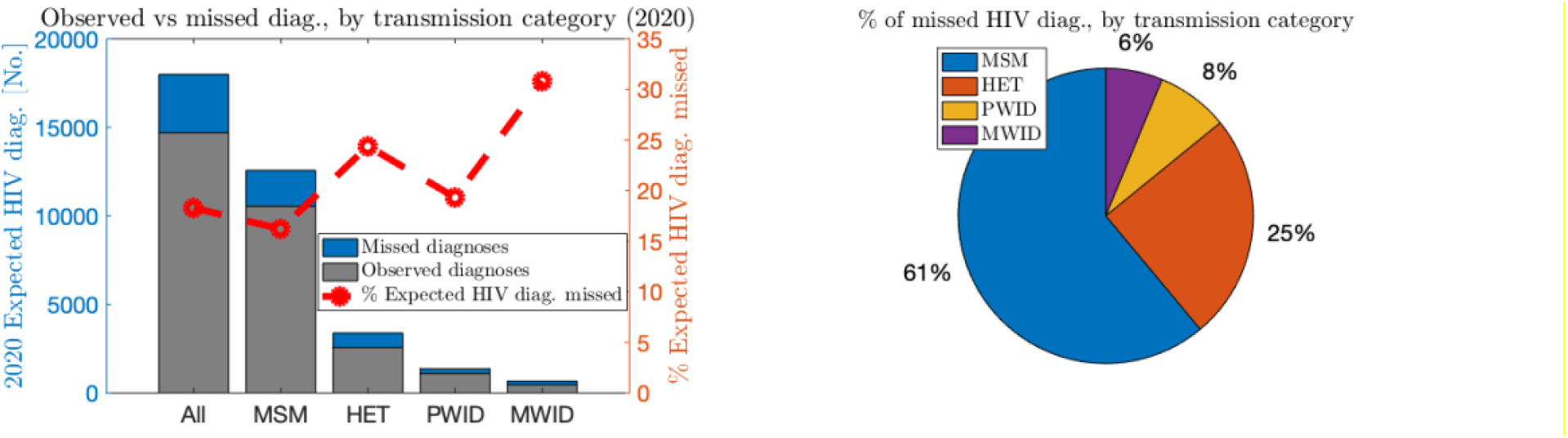
Level of excess missed HIV diagnoses in 2020, by transmission group, United States.

Figure 3 (b) shows the percentage of total excess missed diagnoses in 2020 by transmission group. MSM accounted for 61% of the 3288 excess missed HIV diagnoses in 2020, followed by HET (25%), PWID (8%), and MWID (6%).

In Appendix B, we provide a complete analysis of excess missed diagnoses in 2020, including stratifications by sex assigned at birth, race/ethnicity, and region. Our analysis showed notably elevated levels of missed diagnosis among female sex assigned at birth (∼23-25%), Hispanic/Latino PWH (∼22-25%). In addition, the largest number of missed HIV diagnoses was among black PWH (1,267-1,366), representing roughly 40% of the total missed diagnoses. Differences in missed diagnoses between regions were less pronounced, but appear slightly elevated in the Northeast (∼19-20%) and slightly lower in the Midwest (15%). We also performed stratification robustness analyses, which confirmed the consistency and reliability of our estimates.

In Appendix C, we performed a validation study, using the diagnosis-delay and incidence-based methods to estimate new diagnoses in 2019 among PWH infected from 2010-2018. These estimates showed good agreement with the observed diagnosis data (NHSS data), confirming the accuracy of the methods. Additional analyses confirmed the robustness of the different methods to population stratification.

## Discussion

We developed three distinct methods (one using two formulations) to estimate excess missed diagnoses among PWH infected from 2010-2019. All methods estimate that there were approximately 3100-3300 fewer diagnoses than expected among this population in 2020. Our estimation techniques, despite their differing formulations, are consistently in qualitative and quantitative agreement, providing evidence towards their reliability. These missed diagnoses cannot be attributed to changes in incidence, and firmly establish that altered testing behavior during the COVID-19 pandemic is responsible for a large portion of the observed drop in HIV diagnoses in the United States in 2020. They further indicate COVID-19 and associated disruptions have led to a decrease in awareness of infection status.

Our results indicate that different subpopulations were not equally affected by COVID-19 related disruptions, and the levels of missed diagnosis varied across groups. We found that heterosexuals, PWID, and particularly MWID missed diagnoses at elevated levels. That is, greater percentages of expected HIV diagnoses in 2020 were missed in these groups as compared to MSM. Our further analyses (reported in Appendix B) indicate that females (sex assigned at birth) and Hispanic/Latino PWH also showed disproportionately high levels of missed diagnosis. Despite MSM having a lower proportion of missed diagnoses than other transmission groups, MSM comprise the majority (61%, n∼2,000) of all missed diagnoses (n=3,288).

We note that the true number of excess missed diagnoses of PWH infected pre-pandemic is almost certainly higher than that reported here; due to limitations on completeness of data, our estimates only go back to infections dating from 2010 and later. Considering the decreasing trend in excess missed diagnoses for older infections, as well as the increased probability of a stage 3 (AIDS) disease classification in 11+ years after infection [20], we believe the numbers of excess missed diagnoses from 2009 and earlier are likely smaller compared to those in the years considered. Nonetheless, they are almost certainly non-zero. The numbers shown here should therefore be interpreted as *low-end estimates* for total excess missed diagnoses among PWH infected before 2020.

While the different methods generally agree, we comment on a notable qualitative difference: the incidence-based and diagnosis-delay based methods identified infection-year trends in missed diagnoses among PWID and heterosexuals. These trends were not captured by the diagnosis-based method, which instead suggested a more regular infection-year distribution. However, this is likely due to the construction of the diagnosis-based method, which postulates that undiagnosed infections follow an infection year-distribution based on that of PWH with diagnosed infection. This assumption may not always be appropriate. In contrast, the diagnosis delay and incidence-based models make no assumptions on infection year distribution, and hence may be more suitable for identifying such qualitative trends. We note additionally that in the validation study (provided in Appendix C), the incidence and diagnosis delay-based methods accurately predicted the infection-year distribution of PWID with infection diagnosed in 2019. Thus, we believe that the diagnosis delay and incidence-based methods are more trustworthy in this case, and this trend is likely real. This is an important aspect to consider in future applications of the presented methods.

The public health ramifications of these excess missed diagnoses are potentially severe, as reduced infection awareness is associated with longer delays in linking-to-care and viral suppression and increased HIV transmission [21]–[23]. Increasing testing efforts and innovative strategies towards correcting this diagnosis gap in the coming years may be necessary to avoid putting the goals of the Ending the HIV Epidemic in the United States (EHE) initiative at risk [24]. We showed that our methods may also be applied to population stratifications, to identify groups who missed diagnoses at high levels. Notably, we found that heterosexuals, PWID, MWID, females (sex assigned at birth), and Hispanic/Latino PWH were significantly more likely to have missed their HIV diagnosis in 2020.

Given the potential consequences of this diagnosis gap, future analyses in this area are important. Applying these methods to identify additional disproportionately affected subpopulations may help identify where more targeted or tailored approaches are needed to reach those undiagnosed PWH. Such analyses will help inform efforts towards achieving health equity, eliminating disparities, and improving the health of all population groups. While only one-way stratifications were considered in the present, analyses on further stratification may also be useful in identifying priority populations. Similar projections may also be performed at different geographic scales, including, for instance, individual EHE jurisdictions, to help prioritize intervention strategies. Quantifying the costs and benefits of such interventions, including future increased incidence due to a persisting diagnosis gap, is also critical.

This analysis helps firmly quantify the role of changes in testing behavior in the 2020 pandemic-related diagnosis drop. The results shown here strongly suggest that factors other than reductions in incidence, such as possible disruptions in the provision of testing services and reductions in testing behaviors, resulted in a significant reduction in diagnosis levels. As such, this analysis and the introduced techniques may be further used to help understand and ultimately estimate the other presumed major contributor to the diagnosis drop – the effect of the COVID-19 pandemic on HIV incidence.

## Data Availability

All data produced in the present study are available upon reasonable request to the authors.

## Appendix A

### Full methodological details

#### Diagnosis-based method

Let *D*(*y*) be the number of HIV cases diagnosed in year *y, y* ≤ 2020. To estimate the reduced number of HIV cases diagnosed in 2020 due to the COVID pandemic, we estimate the expected number of HIV diagnoses in 2020 based on the diagnosis trend in recent years prior to 2020. A 4-year log linear regression was used as a best fitting model among models considered based on the Bayesian Information Criterion [25].

Let *E*_*TOT*_(2020) be the total expected number of HIV diagnoses in 2020 and *D*_*TOT*_(2020) the total observed diagnoses number in 2020. Then the total excess missed diagnoses in 2020 *M*_*TOT*_ are given as:

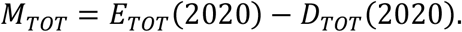

In 2020, this amounted to:

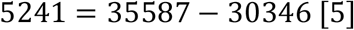

Among those 30346 HIV cases diagnosed in 2020, the year of infection can be estimated based on their CD4 test dates and results.

Let *D*_*y*_(2020) be the number of diagnoses in 2020 among PWH infected in year *y*. Assuming the distribution by year of infection between diagnosed infections and the excess missed diagnoses in 2020 was the same, the number expected diagnoses reads:

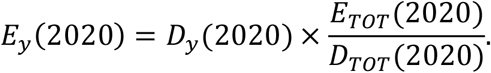

The number of excess missed diagnoses in 2020 among PWH infected in a given year *y, M*_*y*_, is then obtained as:

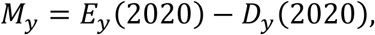

which is then calculated for all relevant infection years.

However, the above assumption may not be appropriate, as HIV can only be diagnosed post-infection. This means that, while infections that occurred before the current calendar year may be diagnosed at any point during the year, infections that occurred during the current year will have only six months, on average, to be diagnosed. Considering this fact, we reduce the estimated number of excess missed diagnoses among PWH infected in 2020 by 50%:

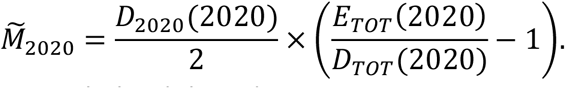

Figure A1 depicts the reasoning behind this adjustment.

However, even if 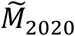 is changed, this does not affect the value of *E*_*TOT*_(2020) and hence our estimate of excess missed diagnoses across all infection years *M*_*TOT*_ has not changed. Since excess missed diagnoses in 2020 have been halved, we must account for the other half (equal to 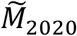) among the other remaining infection years. This is done proportionally, based on the number of (pre-adjustment) excess missed diagnoses in each infection year *y, y* ≤ 2019, and denoted 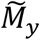:

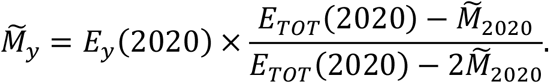

**Figure A1:**
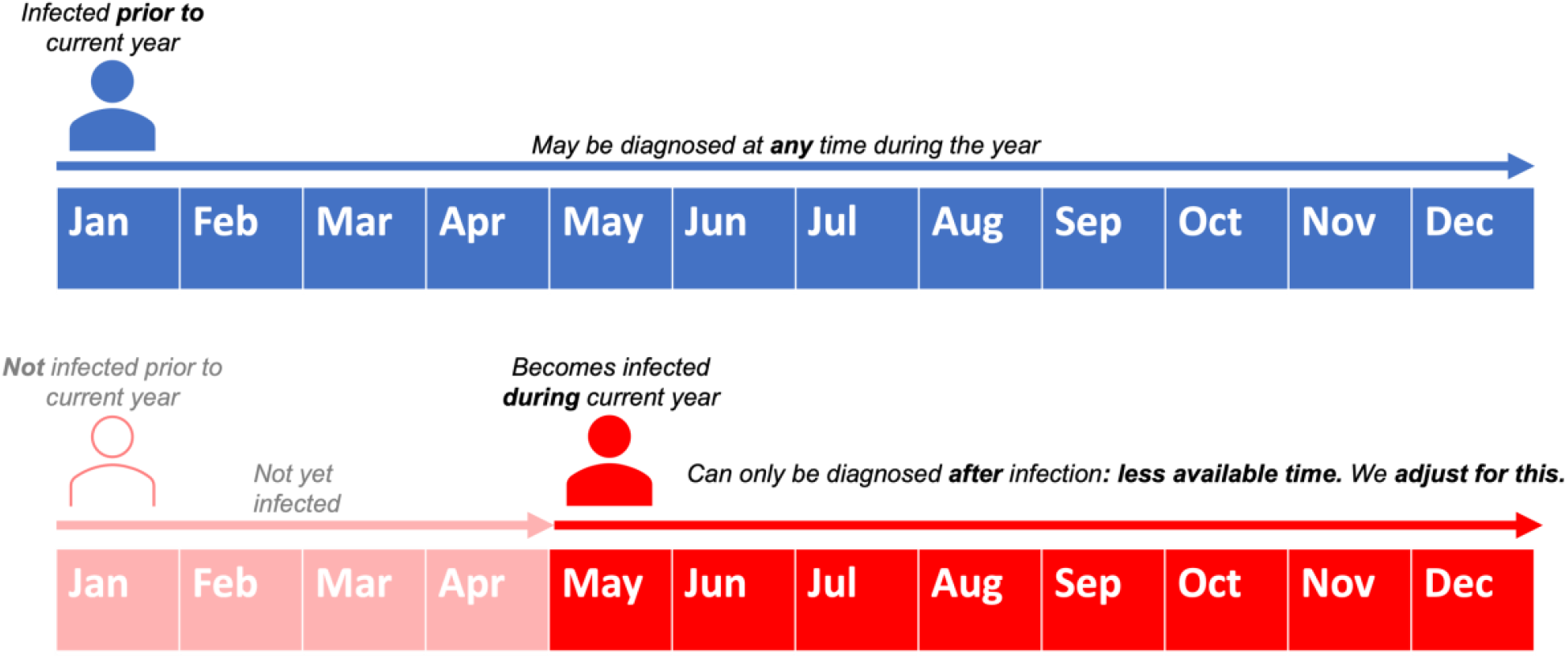
Figure depicting the need to incorporate an adjustment in the diagnosis-based method

**Table A1:** Example calculation for diagnosis-based method.

|  | Year of Infection (y) |  |  |  |  |  |  |  |  |  |  |  |  | Total |
| --- | --- | --- | --- | --- | --- | --- | --- | --- | --- | --- | --- | --- | --- | --- |
|  | <=2008 | 2009 | 2010 | 2011 | 2012 | 2013 | 2014 | 2015 | 2016 | 2017 | 2018 | 2019 | 2020 |  |
| $D_y(2020)$ | 5136 | 608 | 785 | 1116 | 1208 | 1334 | 1552 | 1629 | 1731 | 1781 | 1803 | 1757 | 9907 | 30346 |
| $E_y(2020)$ | 6023 | 713 | 921 | 1309 | 1417 | 1564 | 1820 | 1910 | 2030 | 2089 | 2114 | 2060 | 11618 | 35587 |
| $M_y$ | 887 | 105 | 136 | 193 | 209 | 230 | 268 | 281 | 299 | 308 | 311 | 303 | 1711 | 5241 |
| $\tilde{M}_y$ | 1102 | 130 | 169 | 240 | 260 | 286 | 333 | 349 | 371 | 383 | 386 | 376 | 856 | 5241 |

See example calculation for the overall total in Table A1.

#### Incidence-based method

Denote the number of *infections* in a year *y* as

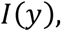

the number *diagnoses* in year *x* of individuals *infected* in year *y* as:

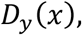

and the probability in year *x* that an individual will receive a diagnosis *k* years after infection as:

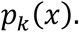

By construction, these obey the following relation:

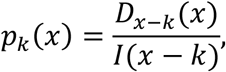

the probability that an, in year *x*, an individual will receive a diagnosis *k* years after infection (or, equivalently, the probability that someone infected in year *x* − *k* will receive a diagnosis in year *x*).

The following are available from surveillance data:

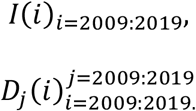

The *I*(*i*) are available from incidence estimates published by the Centers for Disease Control and Prevention [15]–[17]. The *D*_*j*_(*i*) are available from the National HIV Surveillance System, and are the same data used in estimating HIV incidence [7]–[9], [15]–[17].

From the *I*(*i*) and *D*_*j*_(*i*), we compute the following:

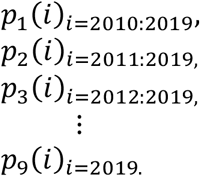

Using these *p*_*k*_(*i*), we then fit functions 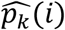, that allow us to estimate 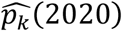 for *k* = 1: 10. Then the *expected diagnoses* in 2020 of persons infected in e.g., 2018, *E*_2018_(2020)is given by:

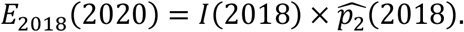

The corresponding excess missed diagnoses *M*_2018_(2020) are then computed as:

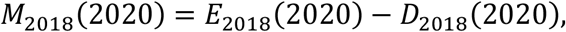

with total excess missed diagnoses *M*_*TOT*_:

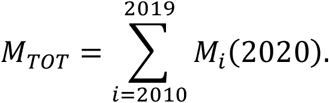

All that remains are the specific definitions of the 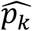. In theory, any such function designed to interpolating *p*_*k*_ in a reasonable way is acceptable. In practice, we acknowledge that a specific choice of interpolation method may depend on the available data, its reliability, and its relevance to current conditions.

In the present work, we considered two definitions of 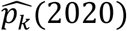, one in which we used a recent-year linear regression, and one in which we considered 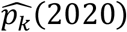 as the three year average of 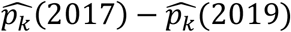^2^. We depict the different 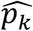 in Figs. A2 (linear) and A3 (average).

**Figure A2:**
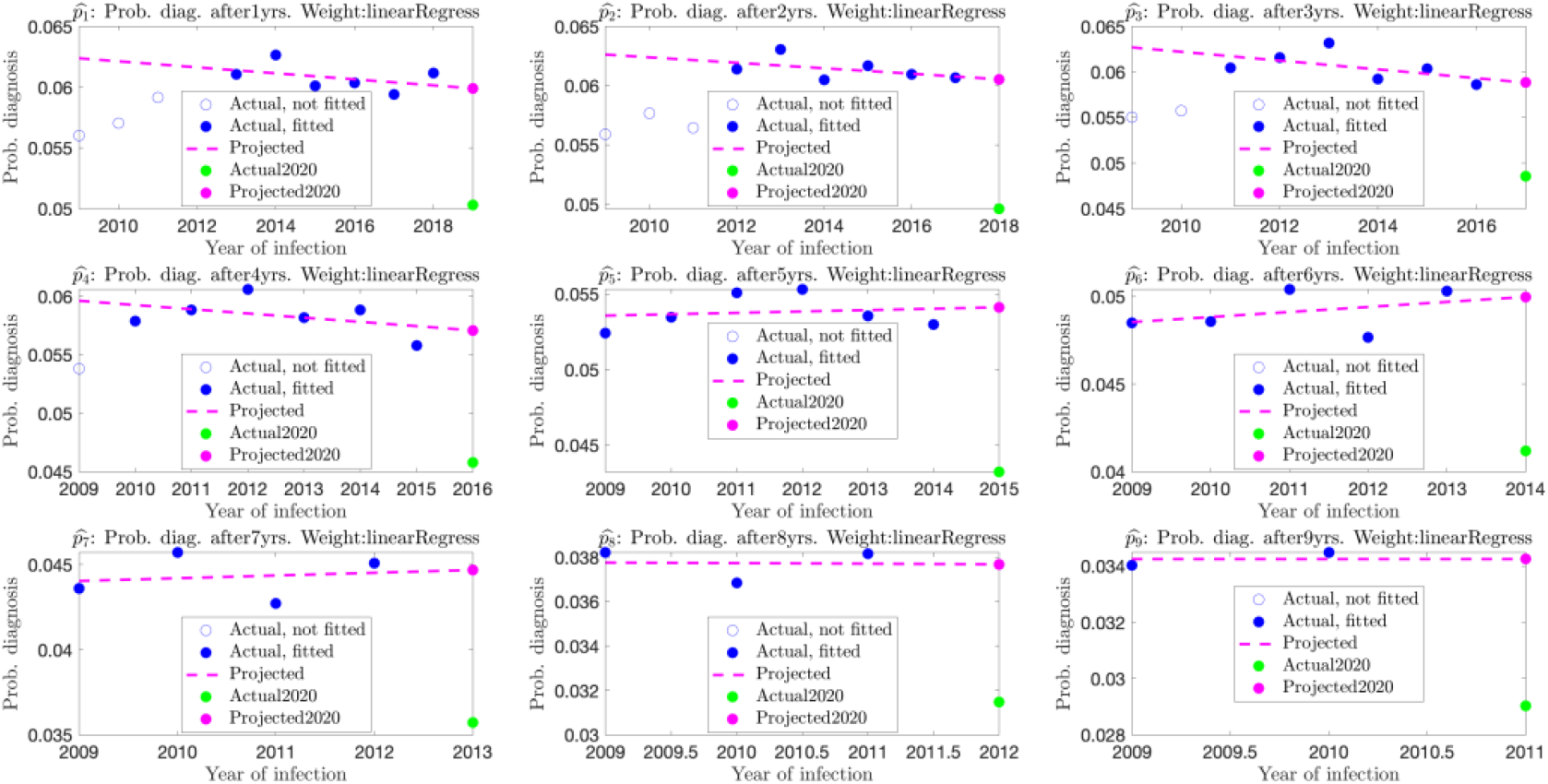
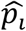 *as defined by linear regression*.

As our results (main text, Appendix B-C) show, the two interpolation methods give similar results. In theory, an extrapolation (such as the linear regression) may better capture a trend, while an average may be less sensitive to noisy data. However, given the similar results obtained, these advantages and disadvantages remain mostly theoretical in the scope of the current analysis.

**Figure A3:**
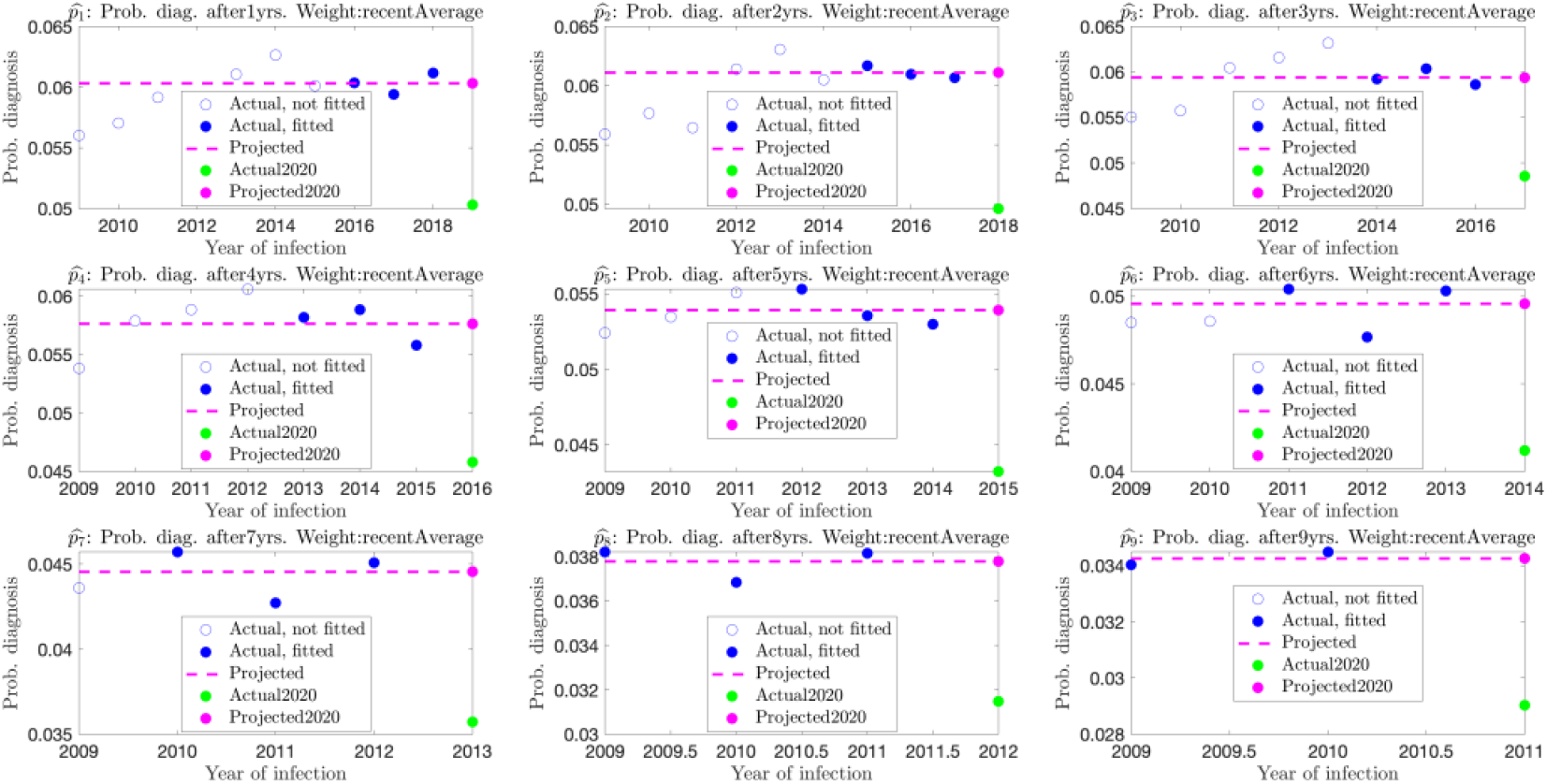
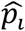 *as defined by a recent-year average*.

#### Diagnosis delay-based method

As mentioned in the methods section, this method is structurally similar to the incidence-based method. The major difference of the two methods is how to estimate the diagnose delay probabilities. This method starts from estimating the cumulative probability that someone infected with HIV will receive a diagnosis within y years after infection. These probabilities are then used to estimate the number of infections and the probability of diagnosis during the *y*-th year after infection.

Let *P*(*k*) be the cumulative probability that someone infected in a calendar year *y* and received a diagnosis during calendar year *y* + *k*. First, we select an *N* and estimate *P*(*N*) using CD4 data from infections diagnosed in recent years. Based on their first CD4 and the CD4 depletion model, we can estimate this probability by the proportion of new diagnoses with an estimated infection date within N years before diagnosis. More specifically,

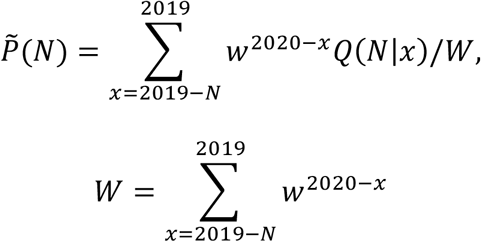

where *w* is a weight (taken as 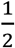 here, see [8]) and *Q*(*N*|*x*) is the proportion of infections diagnosed in year *x* with an estimated infection date within *N* years before diagnosis.

For *k* < *N*, we estimate *P*(*k*) using *P*(*N*) and proportions *P*(*k* − 1|*k*) with the denominator given by the HIV infections diagnosed before 2020 with an estimated infection date before year 2020 − *k* and the numerator given by the number infections diagnosed within *k* − 1 years after infection among those in the denominator:

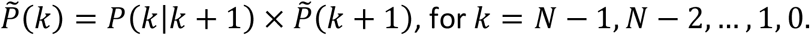

Let *I*(*y*) be the number of infections in year *y, D*(*y*)the number of infections with an estimated infection date in year *y* and with HIV diagnosed during years [*y*, 2019]. Then *I*(*y*) is estimated as:

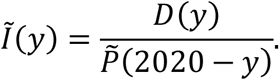

Using the cumulative probabilities P(k), the probability of receiving a diagnosis exactly *k* years after infection, 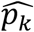, can then simply derived from

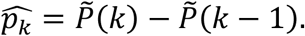

The expected diagnoses for infections in year *y* and diagnosed in 2020 are then given by:

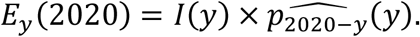

In turn, the excess missed diagnoses in 2020 for PWH are calculated as:

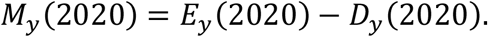

## Appendix B

*Additional analyses of missed diagnoses in 2020*.

### Introduction

In this section, we applied the same methods used in the main body test (diagnosis-based, diagnosis delay-based, incidence-based (linear regression), incidence-based (averaged)) to compute excess missed diagnoses in 2020 for the following stratifications:

1. Sex assigned at birth *(Male, Female)*
2. Region *(Northeast, West, South, Midwest)*
3. Race/ethnicity *(Black or African American, Hispanic/Latino, White)*
4. Transmission group *(MSM, HET, PWID, MWID)*

### Methods

We calculated excess missed diagnoses with each method, for each stratification, in each infection year. We also computed, for each method and each stratification, the percentage of expected diagnoses that were missed.

We also assessed the robustness of our estimation methods to stratification. We refer to the sum of the different population stratifications as *SeparateSum* and the full, population-level analyses (reported in the main text) as *TotalPopulation*. For example, for sex assigned at birth, we have:

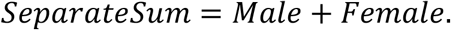

We then evaluated robustness to stratification by comparing *SeparateSum* and *TotalPopulation*. Ideally, the two quantities should be similar – that is, the analyses on the distinct subgroups, when taken together, should resemble the analysis at the entire population level. We quantified stratification robustness by two metrics: *% total difference*, and *% year-by-year difference*.

The *% total difference* is given by the formula:

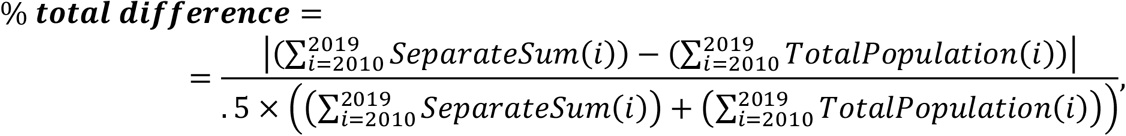

and quantifies the difference in total diagnoses between the summed stratification and full-population analyses. Similarly, the *% year-by-year difference* is given by the formula:

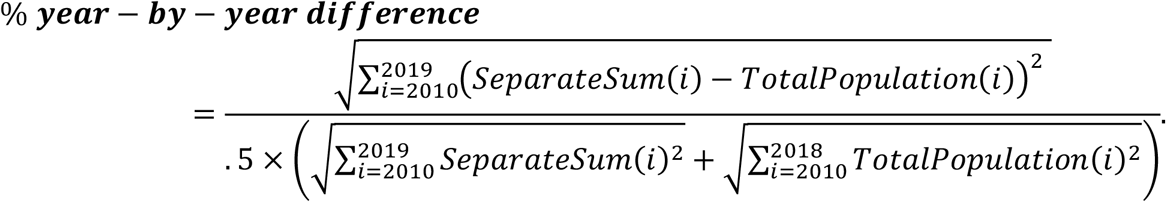

This metric quantifies the agreement in infection-year distribution between the full-population and summed stratification analyses. Note that it is the relative difference in Euclidean Norm over the infection-year period.

### Results

The results are shown in the following plots/tables:

1. Sex assigned at birth *(Male, Female):* Table/Figure B1-B2
2. Region *(Northeast, West, South, Midwest)*: Table/Figure B3-B4
3. Race/ethnicity *(Black or African American, Hispanic/Latino, White)*: Table/Figure B5-B6
4. Transmission group *(MSM, HET, PWID, MWID)*: Table/Figure B7-B8

Females (at birth), while comprising a much smaller raw number of missed diagnoses as compared to males (at birth), missed diagnoses at a substantially higher rate than males in 2020 (Figure/Table B1-B2). This was shown by all estimation methods, with the incidence and diagnosis-delay based methods showing a pronounced skew toward more recent infections.

Missed diagnosis rates were elevated in the northeast compared to other regions. We note that the Midwest showed low rates of missed diagnosis across all methods. The South and West were in line with the national average, as all methods indicated around 18% of diagnoses missed (Figure/Table B3-B4).

All methods indicate that Hispanic/Latino PWH missed diagnoses at notably higher rates compared to Black or African American (hereafter referred to as Black) and White PWH. Black and White PWH did not show a substantial difference in missed diagnosis rates, and were both below the national average (Figure/Table B5-B6)

As discussed in the main text, heterosexuals and MWID showed higher levels of missed diagnosis. The methods are in slight disagreement on PWID; with some showing a slightly elevated rate of missed diagnosis and others a highly elevated rate. We can conclude that missed diagnoses were likely elevated in this group, while acknowledging some uncertainty about the magnitude (Figure/Table B7-B8).

Additionally, all methods were robust to stratification, with % total difference consistently under 1%, and the % year-by-year difference consistently under 5% in all cases (and often much lower).

**Table/Figure B1:**
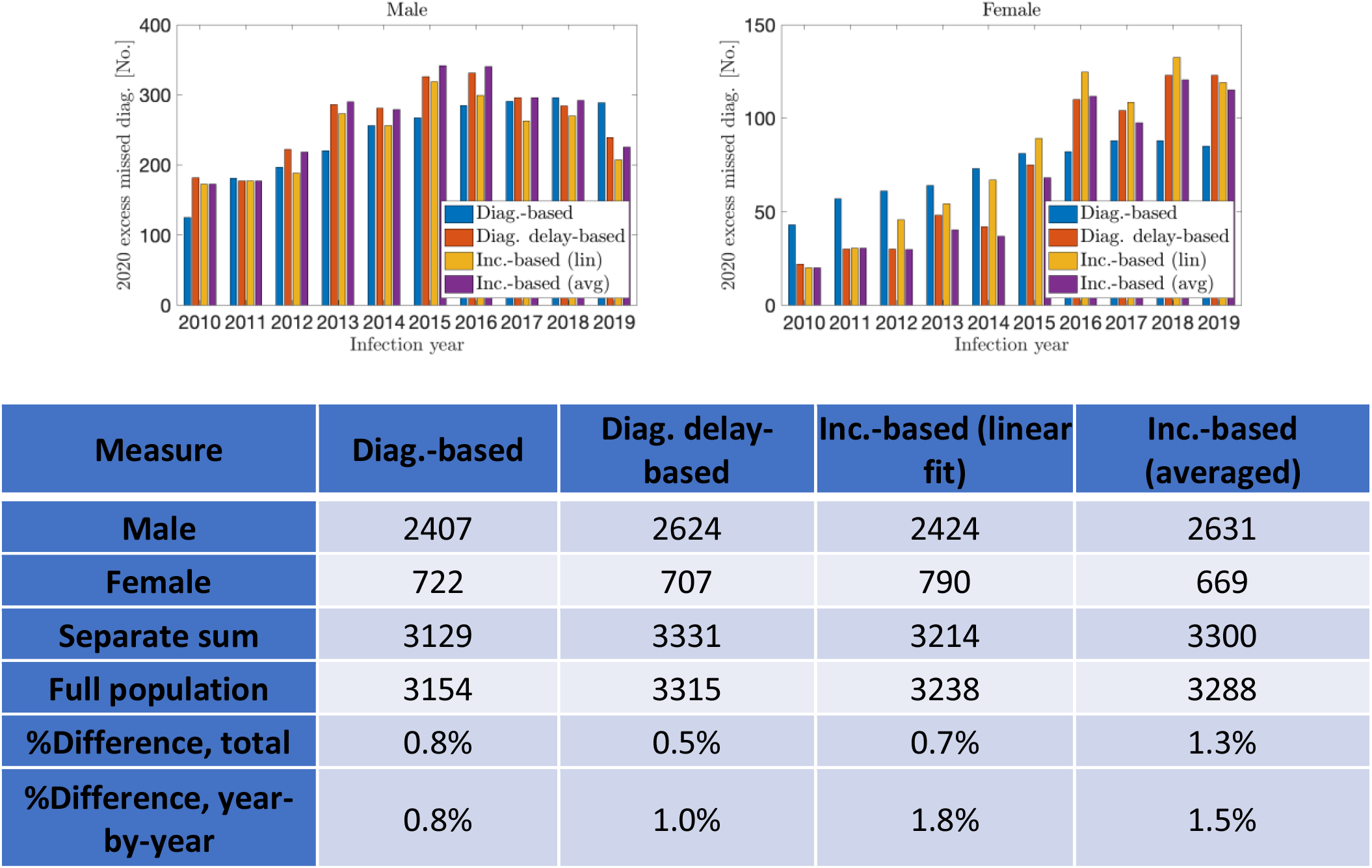
Excess missed HIV diagnoses in 2020, among persons infected during 2010-2019, by sex assigned at birth, United States

**Table/Figure B2:**
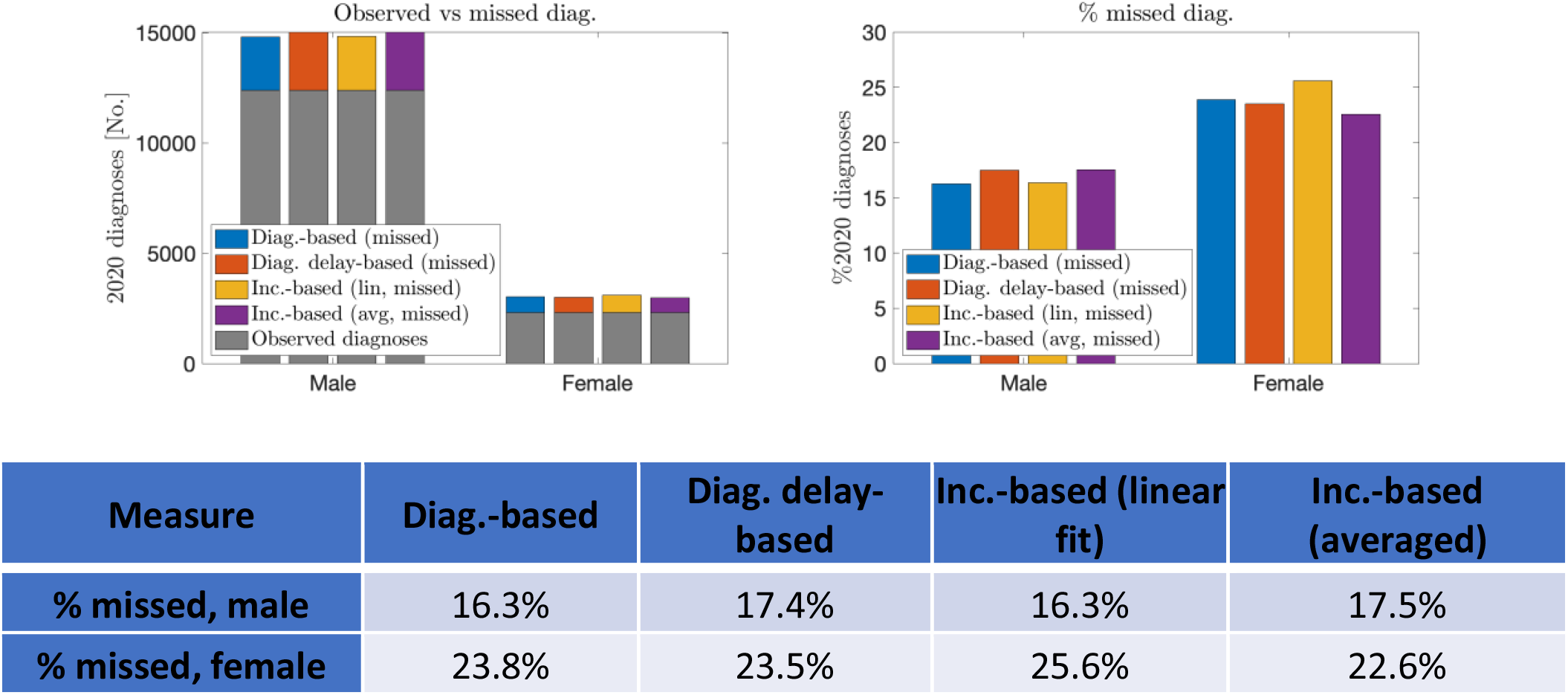
Percent of expected diagnoses missed in 2020, by sex assigned at birth, United States

**Table/Figure B3:**
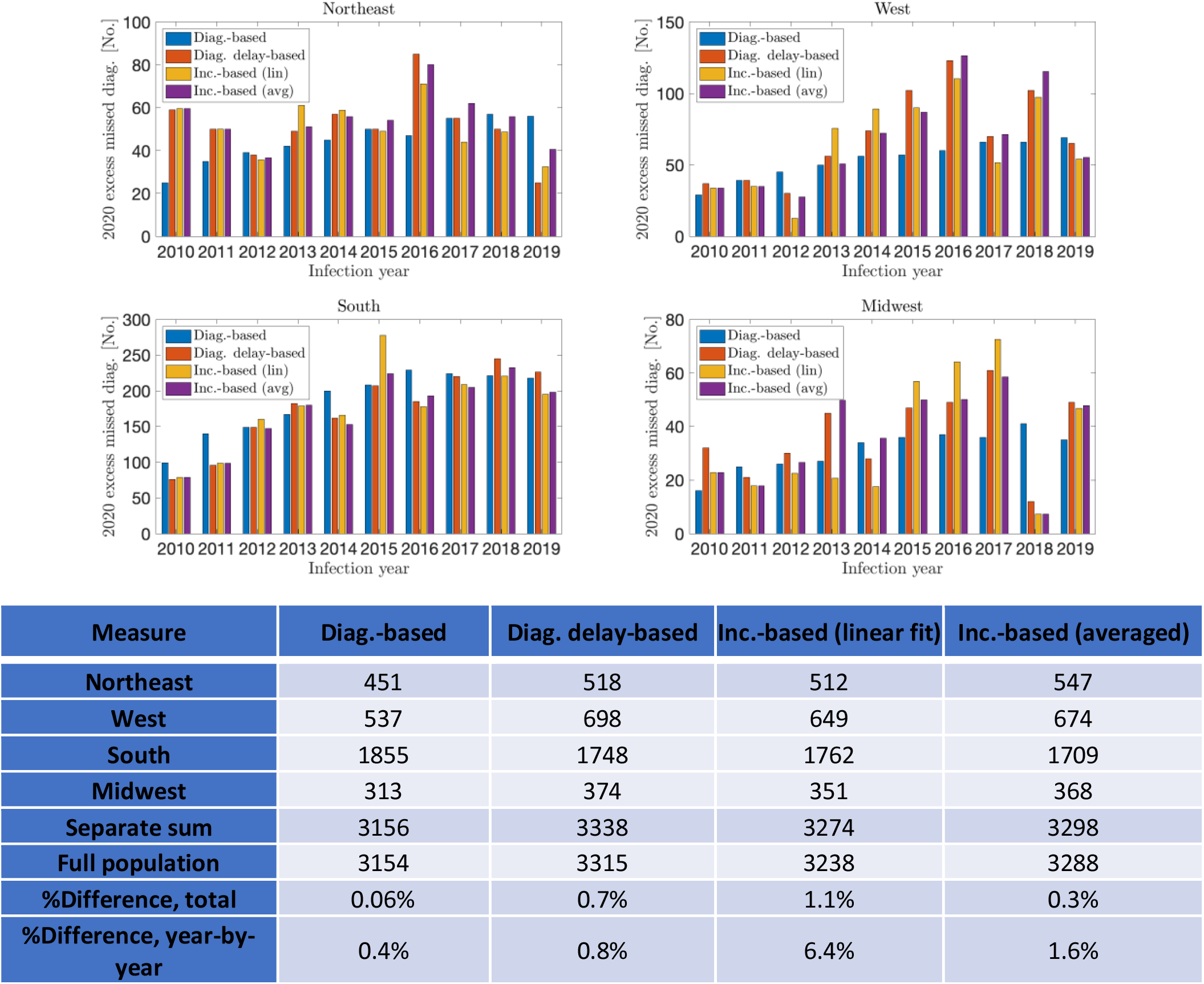
Excess missed HIV diagnoses in 2020, among persons infected during 2010-2019, by region, United States

**Table/Figure B4:**
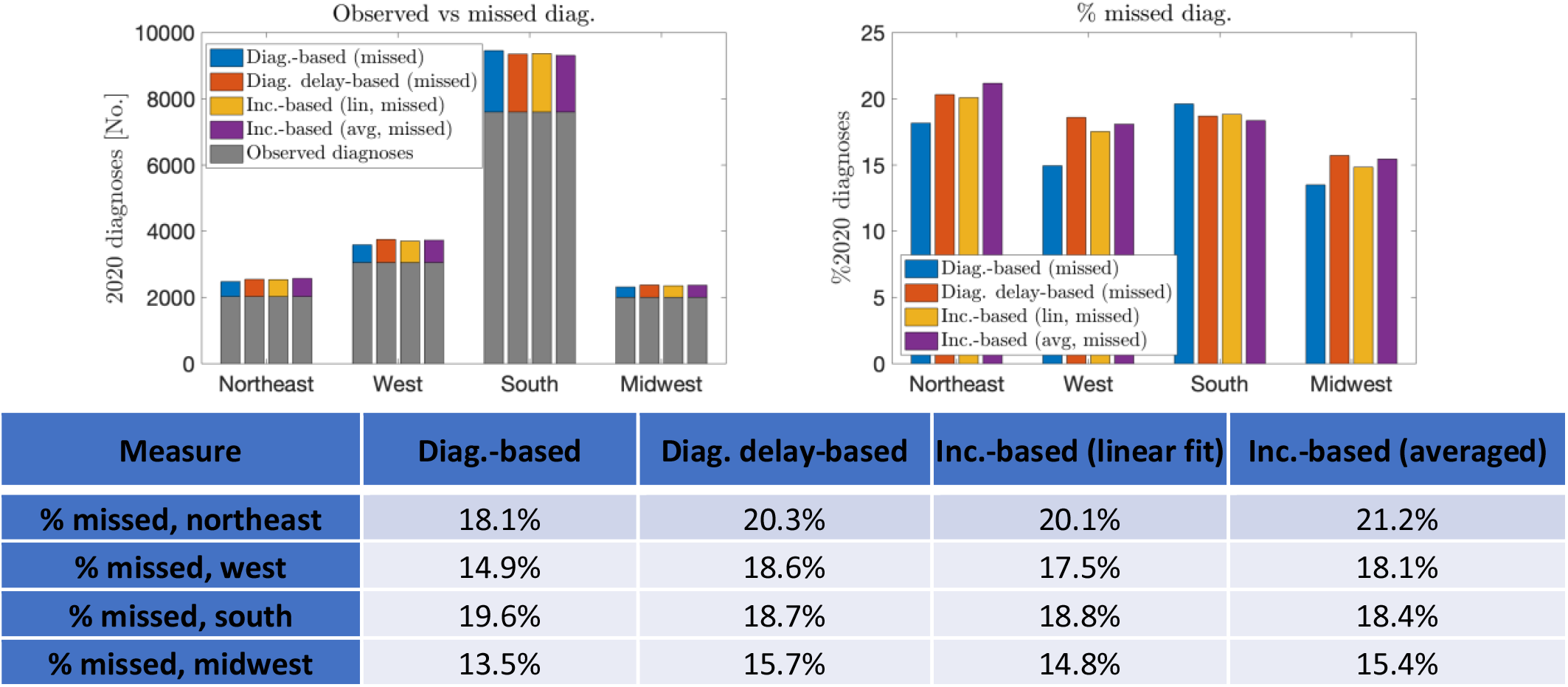
Percent of expected diagnoses missed in 2020, by region, United States

**Table/Figure B5:**
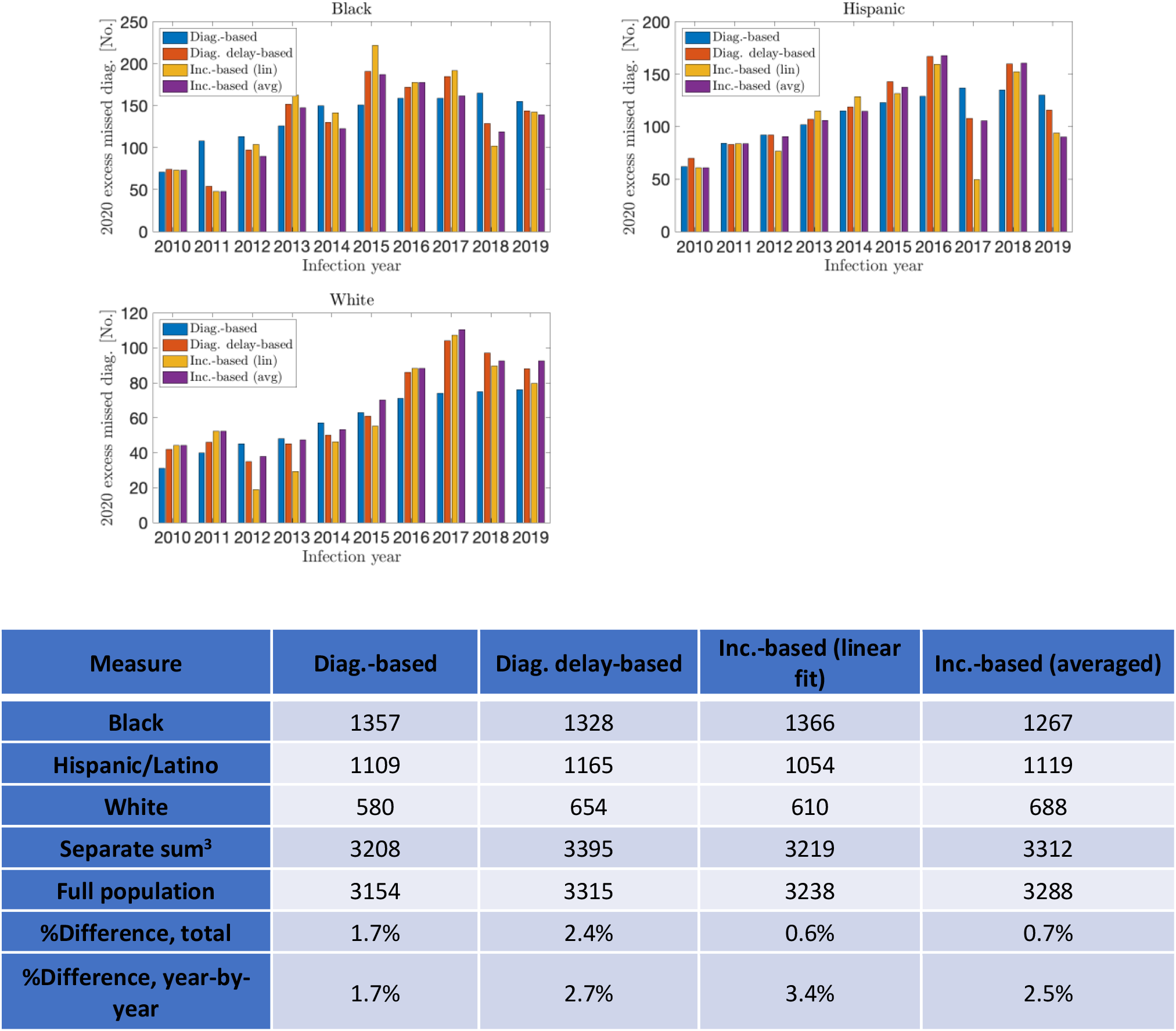
Excess missed HIV diagnoses in 2020, among persons infected during 2010-2019, by race/ethnicity, United States

**Table/Figure B6:**
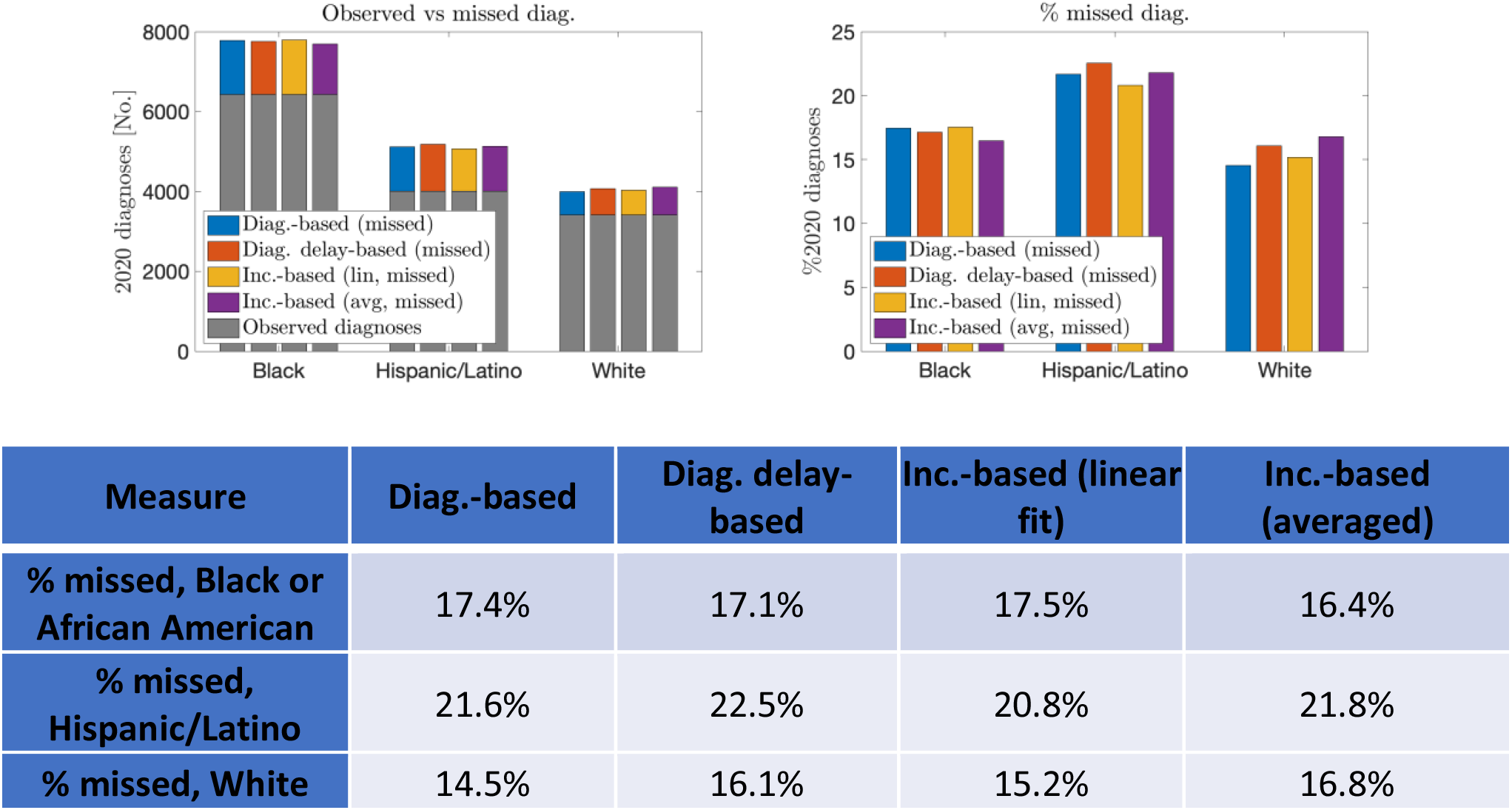
Percent of expected diagnoses missed in 2020, by race/ethnicity, 2010-2019, United States

**Table/Figure B7:**
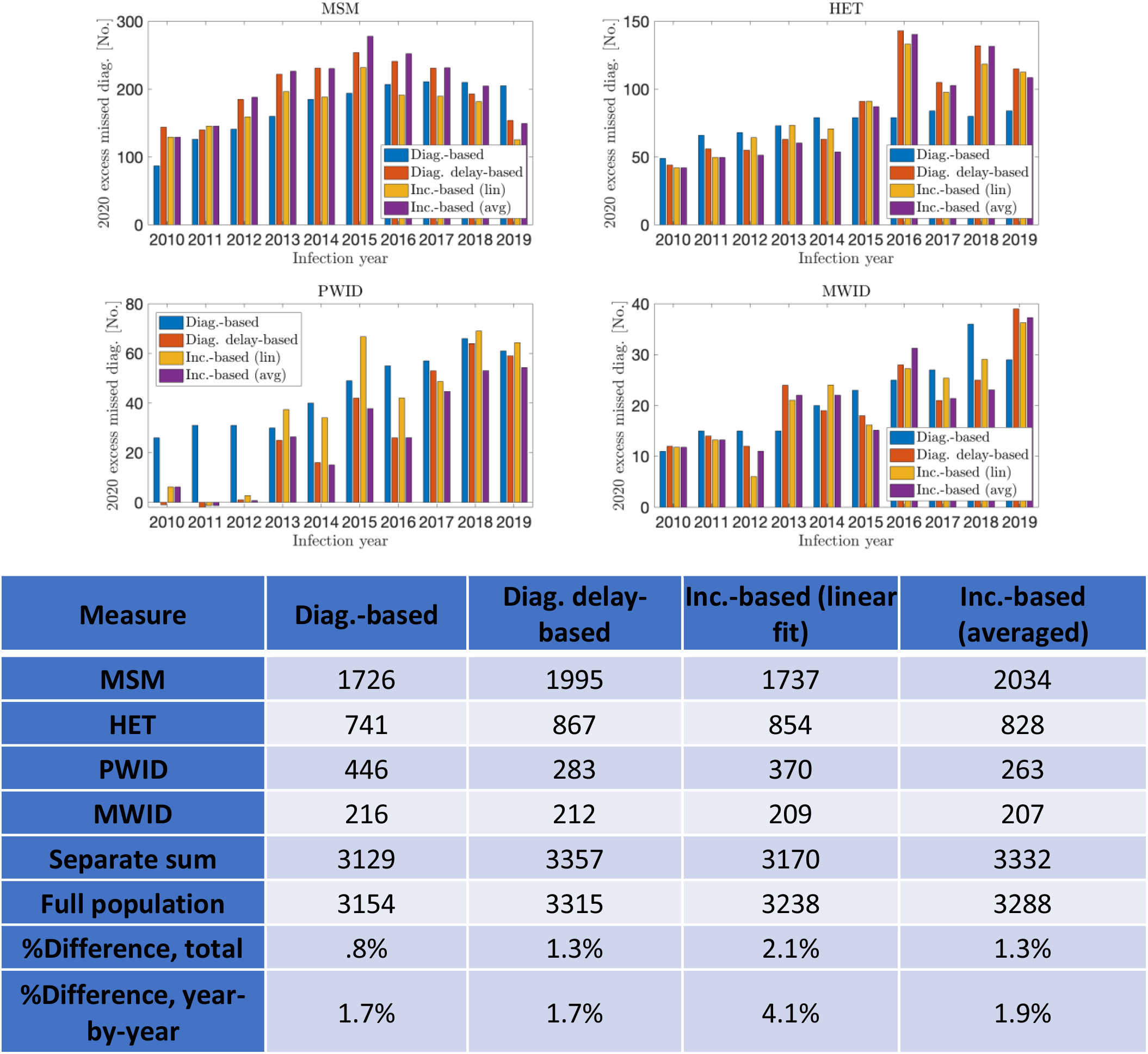
Excess missed HIV diagnoses in 2020, among persons infected during 2010-2019, by transmission group, United States

**Table/Figure B8:**
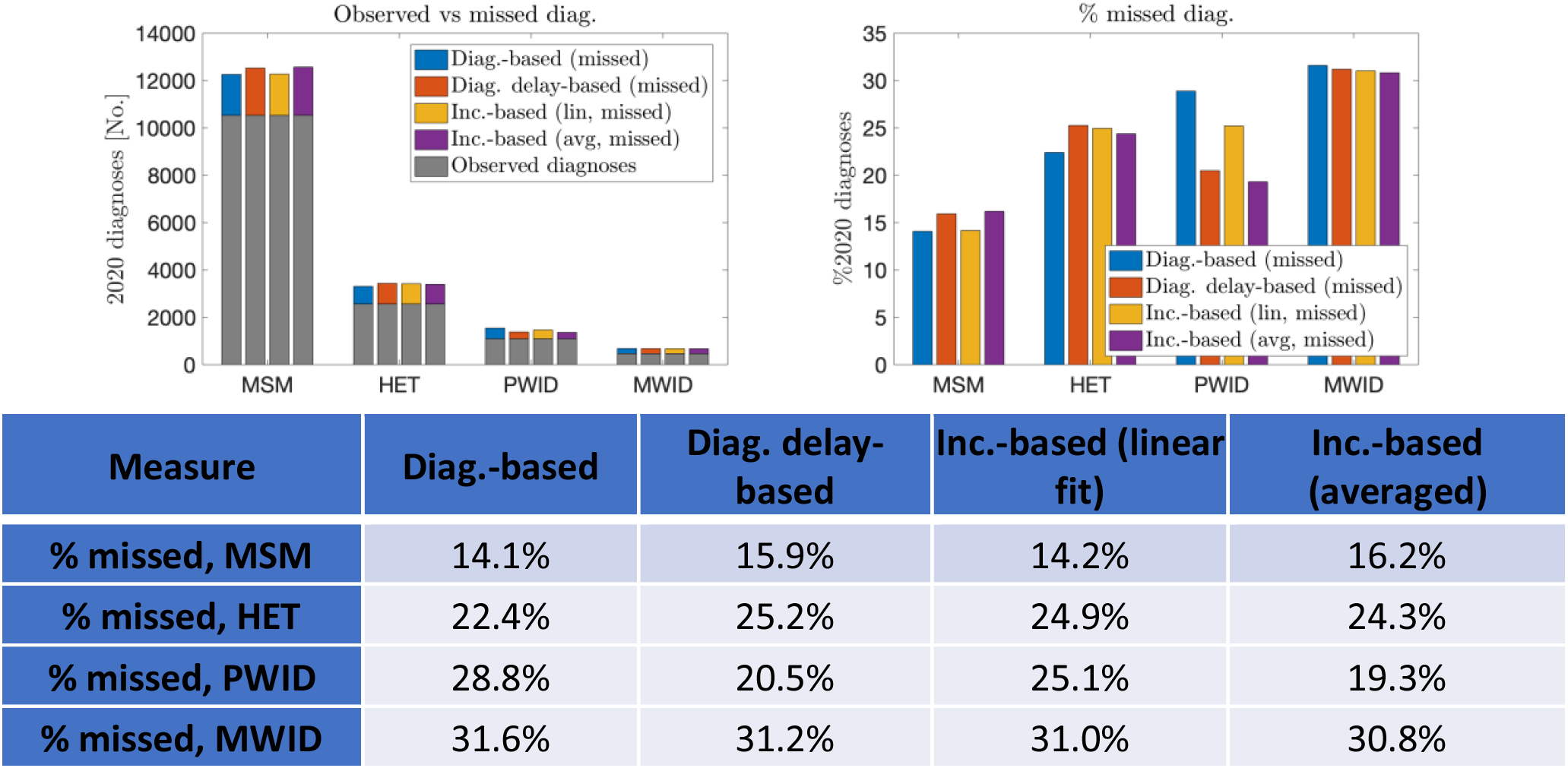
Percent of expected diagnoses missed in 2020, by transmission group, 2010-2019, United States

### Discussion

We analyzed the excess missed diagnoses in 2020 by sex assigned at birth, geographic region, race/ethnicity, and transmission group. The analyses suggest that females (at birth), Hispanic/Latino PWH, heterosexuals, and PWID missed diagnoses at disproportionately high rates. Missed diagnosis rates appear somewhat above average in the northeast and notably below average in the Midwest. While MSM, males (at birth), and Black/White PWH had larger raw numbers of missed diagnosis overall, they showed lower rates of missed diagnosis.

The different methods were largely in agreement, with minor differences. The diagnosis-based method at times gave qualitatively different infection-year trends as compared to the other methods. This is unsurprising, as the other three methods are closely related, in contrast to the diagnosis-based method, which is distinct. Differences in total missed diagnoses, however, remained similar for all methods.

We also studied the robustness of the introduced methods to stratified analyses. All methods were robust to stratification, with the summation of the stratification groups showing little difference when compared to the full population-level analysis.

Based on the results obtained, interventions prioritizing PWID, heterosexuals, females (at birth) and Hispanic/Latino PWH may be particularly important in identifying excess undiagnosed infections among PWH and ensuring prompt linkage to care. These analyses also establish that the introduced methods are well-suited for stratified analyses. Similar analyses may be performed on different population stratifications to help design appropriate interventions.

Further analyses considering additional population stratifications, including two-way (or more) stratifications, should also be performed.

## Appendix C: 2019 Validation

### Introduction

In this appendix, we provide validation for the diagnosis delay-based and incidence-based methods on 2019 diagnosis data. The aim is to demonstrate the validity of our estimation methods by evaluating their performance against known (National HIV Surveillance System, NHSS) data. We do not validate the diagnosis-based method with this approach, as its construction makes it ill-suited for such a task. This is because the diagnosis delay-based method assumes that there is a discrepancy in diagnoses to be explained (see Appendix A). As there is no COVID-19 disruption in this validation study (for the year 2019), applying the diagnosis-based method here does not make sense.

#### Methods

We considered the same NHSS diagnosis data and published incidence data used for our primary study; however, to derive our estimates, we used only diagnosis data of persons over 13 years of age from the years 2010-2018. We then applied the diagnosis delay-based and incidence-based (both linear regression and averaged formulations) methods to project expected diagnoses in 2019 for the infection years 2010-2018. We evaluated the quality of the estimates by comparing the projected and observed diagnoses on 2019 for each infection year.

We applied this analysis to the entire PWH population in the United States, and further considered stratifications by sex assigned at birth, region, race/ethnicity, and transmission group. We evaluated our results both in terms of accuracy compared to measured data and robustness to stratification. We employed several quantitative metrics in our evaluations, and provide the definitions below.

#### Definitions

Note that %*Difference, summed vs. full population* (*total*) and %*Difference, summed vs. full population* (*year by year*) are the same as the % *total difference* and % *year* − *by* − *year difference* introduced in Appendix B, respectively. The terminology is changed here for clarity, as we also considered differences with respect to surveillance data in the present. Familiarity with summation notation is assumed.

- ***%Error (total):*** This is a measure of accuracy, and refers to the difference between the total measured diagnoses in 2019 (across all infection years) and estimated total diagnoses 2019 (across all infection years):

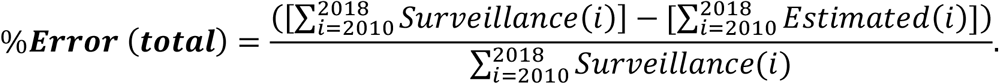
- ***%Error (year-by-year):*** This is a measure of accuracy, and refers to the difference in norm between the measured and estimated diagnoses in 2019 for each infection year, and is expressed as:

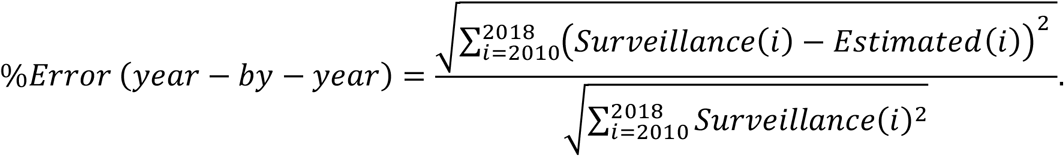
- ***%Difference, summed vs. total:*** This is a measure of robustness to stratification, and refers to the difference in estimated total diagnoses between the analysis on the full population, and the sum of the separate analyses on the stratified groups: %***Difference, summed vs. total***:

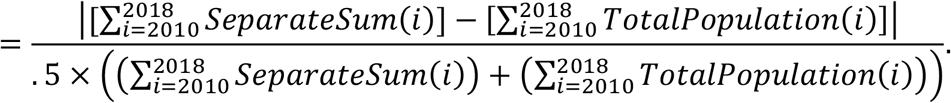
- **%Difference, summed vs. full population (year-by-year):** This is a measure of robustness to stratification, and measures to the difference in infection year-distribution from the estimated diagnoses from the full population analyses, and the sum of the separate stratifications:

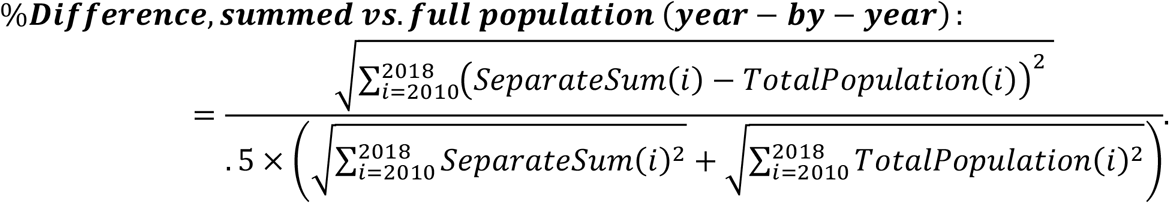

Note that one may apply the triangle inequality with the above metrics to bound the error of the *SeparateSum* against surveillance data on the total population.

### Results

#### Full population analysis

The results are shown in Figure C1 and Table C1. The different methods all accurately projected the diagnoses in 2019 among PWH infected from 2010-2018. Each method estimated the total number of diagnoses within 1% of the surveillance data. The year-by-year results showed similar agreement, with the error in infection year distribution ranging from 3.2-3.6%. These results indicate that the different methods are accurate and provide similar qualitative and quantitative results.

**Figure/Table C1:**
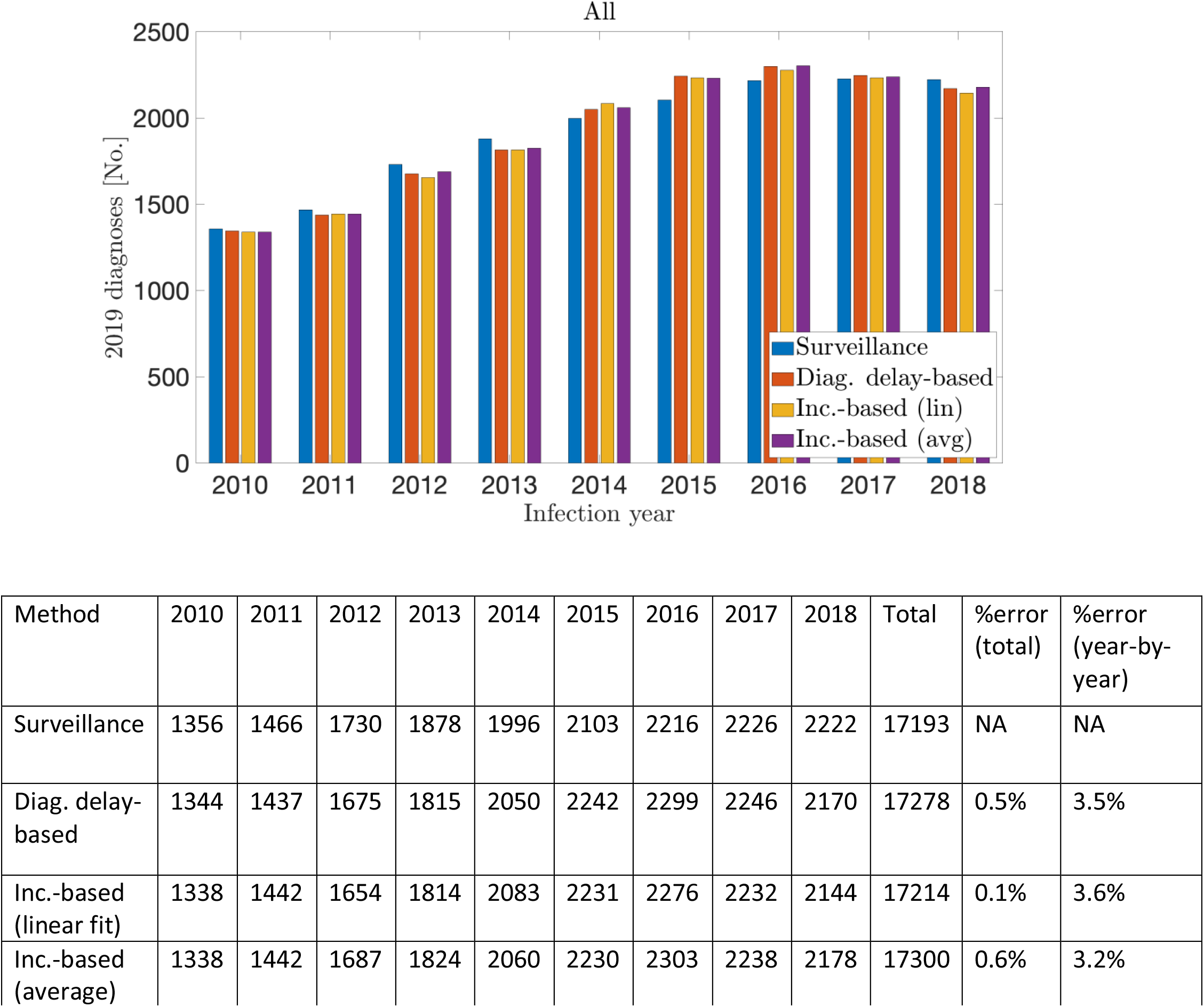
Validation analysis on the full United States PWH population. The different methods agree with both each other and the surveillance data.

#### Stratified analyses

We performed the 2019 validation on the PWH population over the following stratifications:

1. Sex assigned at birth *(Male, Female)*: Table/Figure C2
2. Region *(MSM, HET, PWID, MWID)*: Table/Figure C3
3. Race/ethnicity *(Northeast, West, South, Midwest)*: Table/Figure C4
4. Transmission Group *(Black, Hispanic/Latino, White)*: Table/Figure C5

As with the full population-level analysis, the results were accurate, in terms of both total overall diagnoses and infection year distribution. The overall discrepancy from measured data was nearly always under 5% (and frequently under 1%), and the discrepancy by infection-year nearly always under 10% (and frequently under 5%).

Each method was robust to stratification across all the considered analyses, with the sum of the stratification groups consistently performing similarly to the population-level analyses. The difference between the sum of stratifications and population-level analysis was consistently less than 1% for both overall and year-by-year diagnoses.

**Figure/Table C2:**
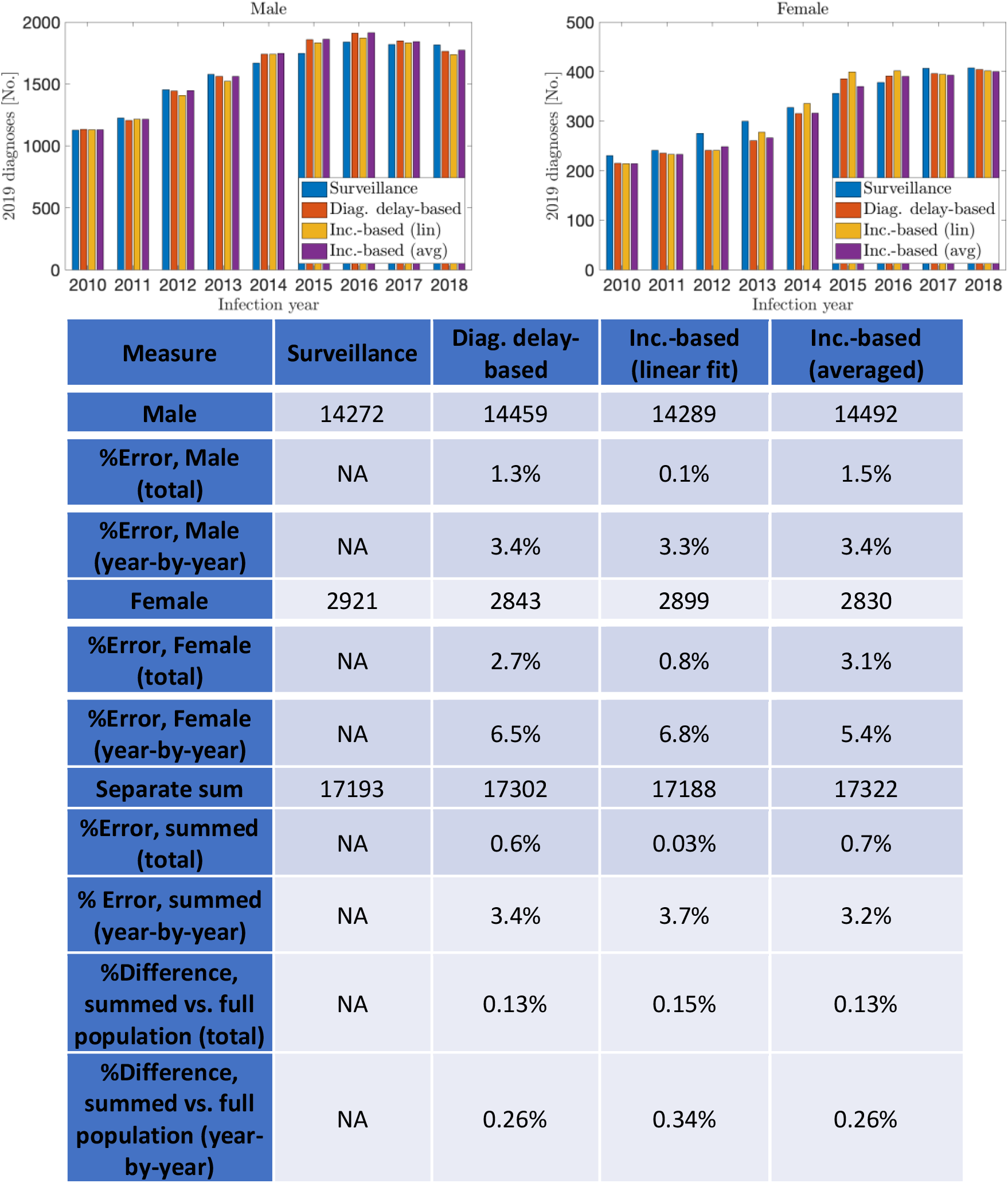
2019 validation, stratified by sex assigned at birth.

**Figure/Table C3:**
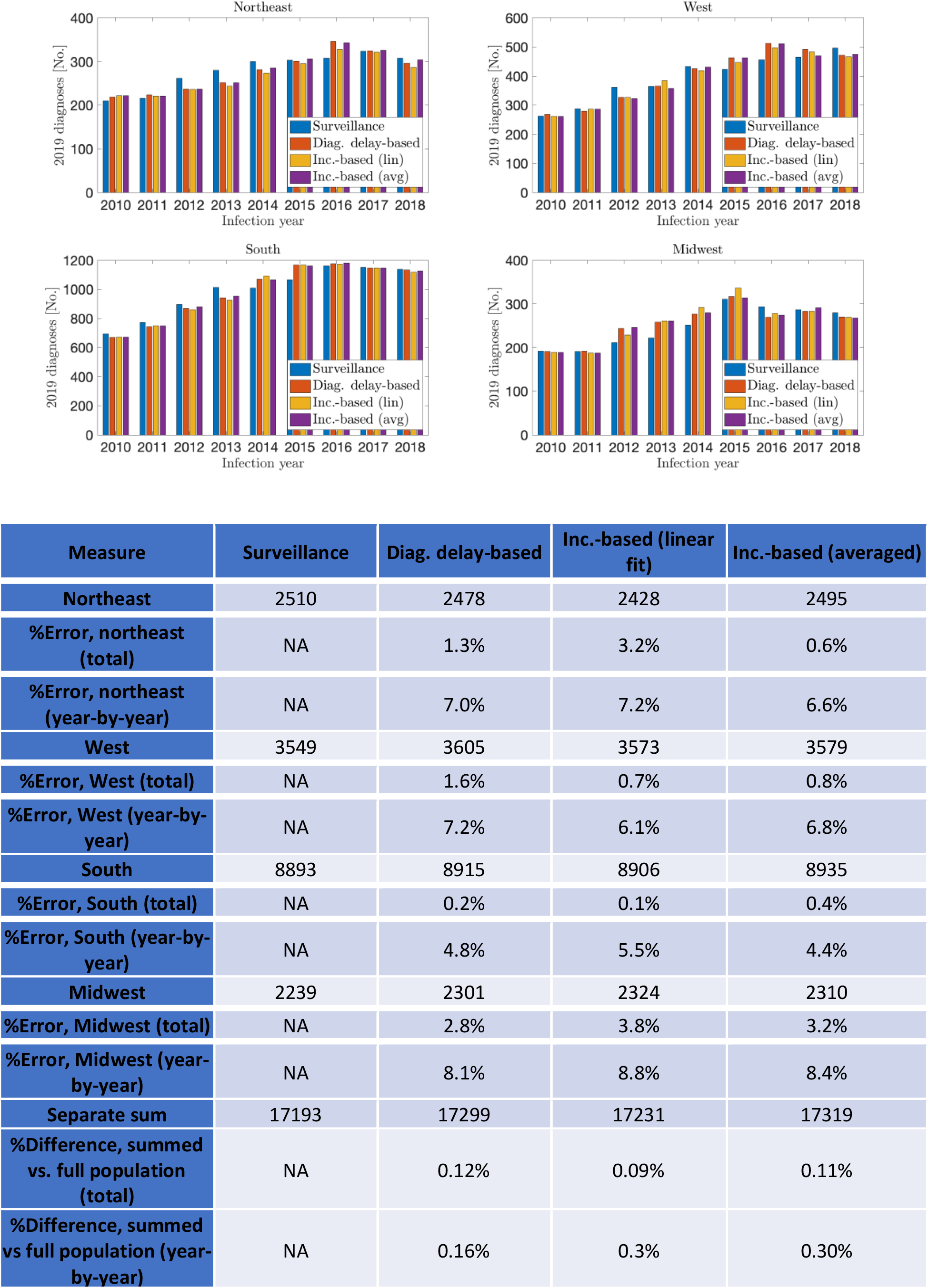
2019 validation, stratified by region

**Figure/Table C4:**
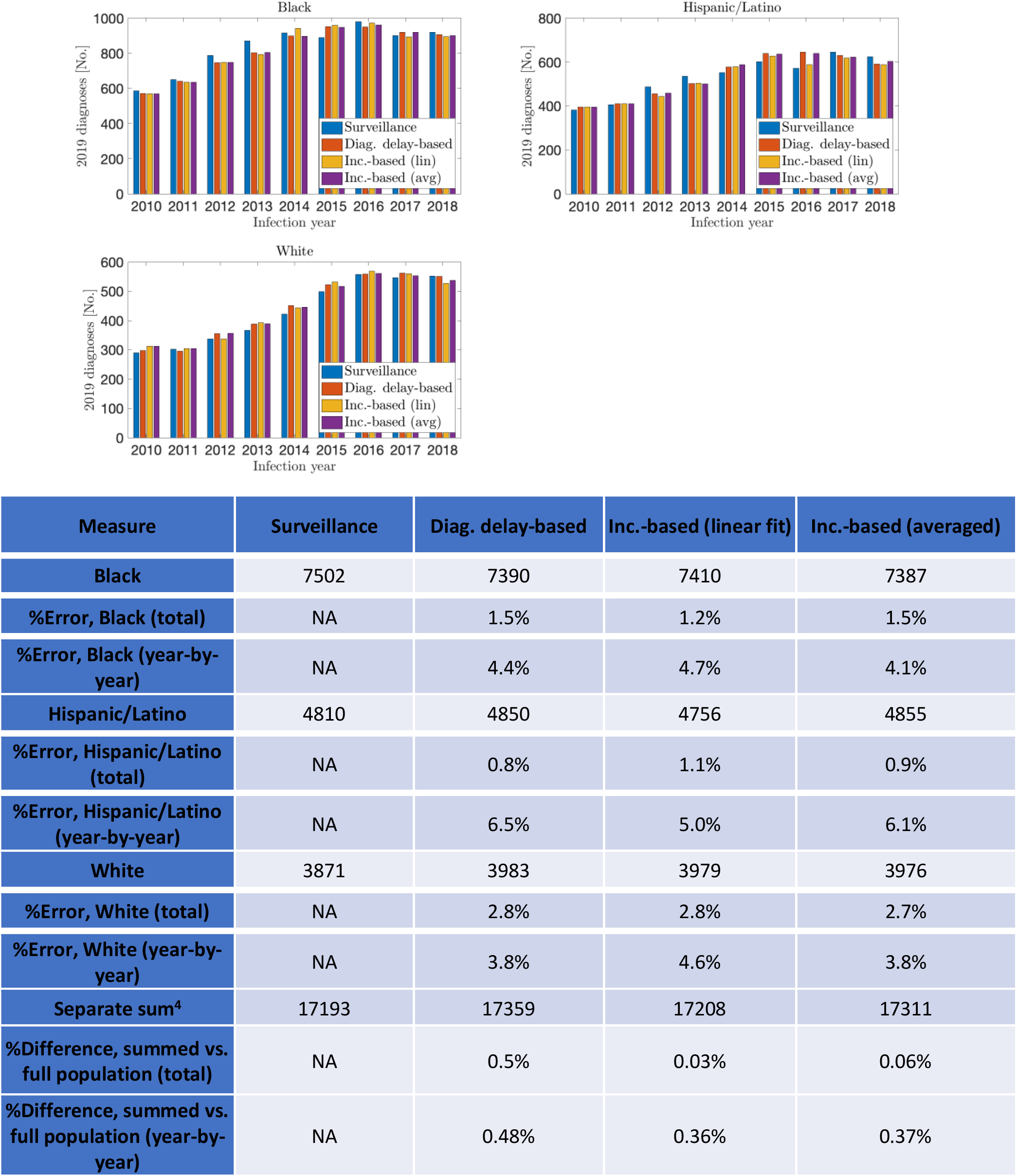
2019 validation, stratified by race/ethnicity, smaller groups aggregated

**Figure/Table C5:**
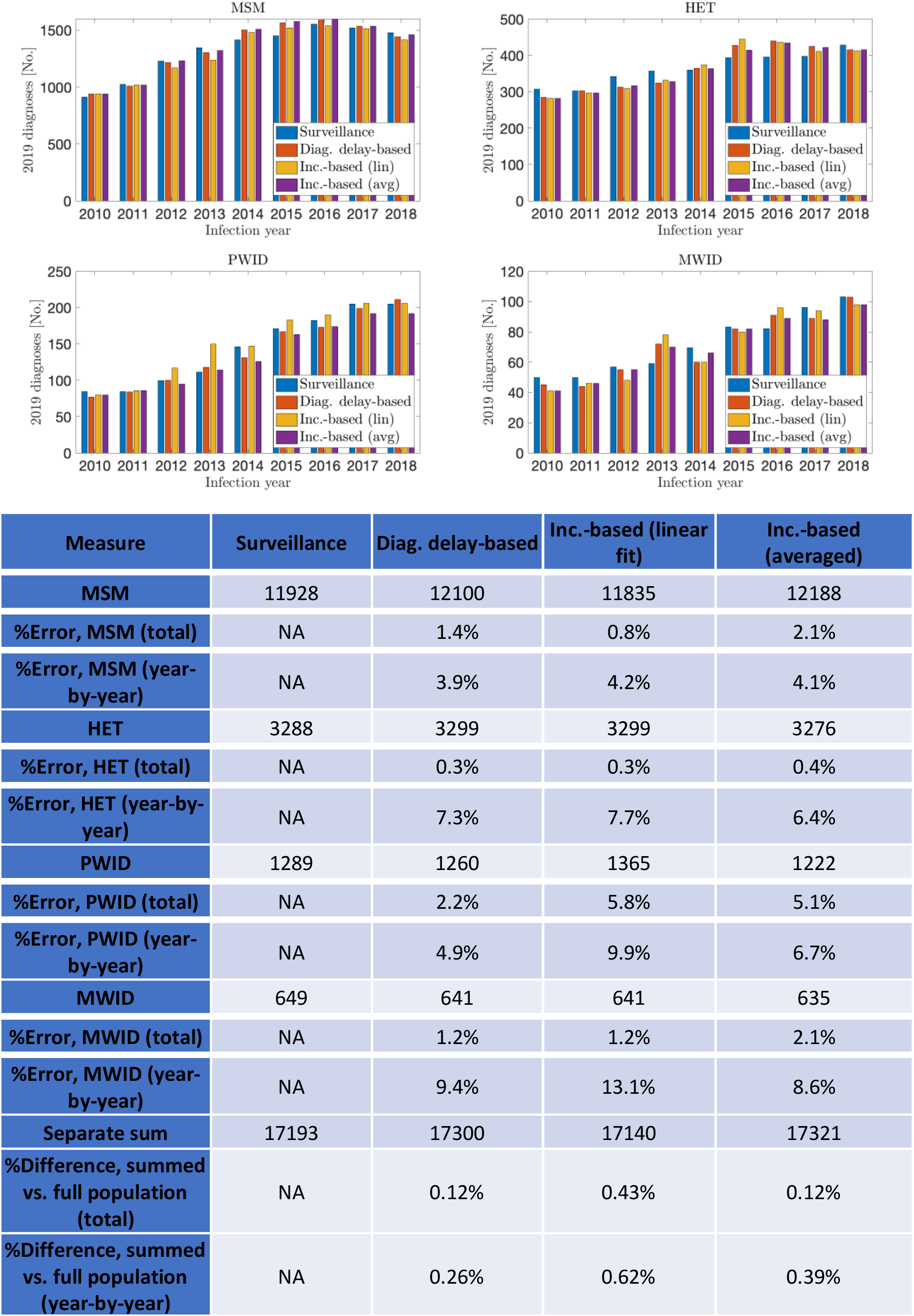
2019 validation, stratified by transmission group.

### Discussion

This validation analysis against 2019 NHSS diagnosis data provides further evidence towards the validity of the introduced diagnosis delay- and incidence-based methods. Each method showed strong agreement with the 2019 surveillance data, with little difference between methods. The results remained accurate when applied to population stratifications, confirming their suitability for such analyses. The summed stratified analyses also differed little when compared to the full population-level analysis, further demonstrating the robustness of the introduced methods to stratification.

In sum, our study on 2019 data provides compelling evidence towards the validity of our methodology, further increasing our confidence in our 2020 estimates and the reliability of future analyses.

## Footnotes

1 Note the method used in this plot is the averaged incidence-based method, similar qualitatively and quantitatively to the diagnosis delay-based method in this analysis. The other methods gave similar results, and the complete analyses are shown in Appendix B.

2 Two- and one-year averages when such years are the only options. Note that for these cases 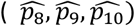, the average was used instead of a linear regression for the “linear regression” evaluations, for obvious reasons. Thus, the two presented interpolation methods do agree on these years.

3 Note that this sum includes smaller racial/ethnic groups, not listed due to small population sizes.

4 Note that this sum includes smaller racial/ethnic groups, not listed due to small population sizes.

## Notes

### Competing Interest Statement

The authors have declared no competing interest.

### Funding Statement

This study did not receive any funding.

### Author Declarations

The data used for this analysis was collected as part of the public health program activity called PS18-1802 Integrated HIV Surveillance and Prevention Programs for Health Departments (Component A), which is a routine disease surveillance activity across 60 jurisdictions throughout the U.S. A project determination made by the National Center for HIV, Hepatitis, STD, and TB Prevention (NCHHSTP) on behalf of The Centers for Disease Control and Prevention (CDC) on January 22, 2018 determined this project to be exempt from needing IRB review as it is not human subjects research. The National Center for HIV, Hepatitis, STD, and TB Prevention (NCHHSTP) on behalf of The Centers for Disease Control and Prevention (CDC) approved the use of this data in the current document on August 25, 2022.

## References

[1] Y.-L. A. Huang, W. Zhu, J. Wiener, A. P. Kourtis, H. I. Hall, and K. W. Hoover, “Impact of Coronavirus Disease 2019 (COVID-19) on Human Immunodeficiency Virus (HIV) Pre-exposure Prophylaxis Prescriptions in the United States—A Time-Series Analysis,” Clinical Infectious Diseases, Jan. 2022, doi: 10.1093/cid/ciac038.

[2] W. Zhu et al., “Impact of the COVID–19 pandemic on prescriptions for antiretroviral drugs for HIV treatment in the United States, 2019–2021,” AIDS, Jul. 2022, doi: 10.1097/QAD.0000000000003315.

[3] E. Moitra et al., “Impact of the COVID-19 pandemic on HIV testing rates across four geographically diverse urban centres in the United States: An observational study,” The Lancet Regional Health - Americas, vol. 7, p. 100159, Mar. 2022, doi: 10.1016/j.lana.2021.100159.

[4] F. Rick, W. Odoke, J. Hombergh, A. S. Benzaken, and V. I. Avelino-Silva, “Impact of coronavirus disease (COVID-19) on HIV testing and care provision across four continents,” HIV Medicine, vol. 23, no. 2, pp. 169–177, Feb. 2022, doi: 10.1111/hiv.13180.

[5] Centers for Disease Control and Prevention, “HIV Surveillance Report, 2020,” vol. 33, 2022. https://www.cdc.gov/hiv/library/reports/hiv-surveillance.html (accessed Jun. 01, 2022).

[6] E. A. DiNenno et al., “HIV Testing Before and During the COVID-19 Pandemic — United States, 2019–2020,” MMWR. Morbidity and Mortality Weekly Report, vol. 71, no. 25, pp. 820–824, Jun. 2022, doi: 10.15585/mmwr.mm7125a2.

[7] S. Singh, R. Song, A. S. Johnson, E. McCray, and H. I. Hall, “HIV Incidence, Prevalence, and Undiagnosed Infections in U.S. Men Who Have Sex With Men,” Annals of Internal Medicine, vol. 168, no. 10, p. 685, May 2018, doi: 10.7326/M17-2082.

[8] Song, H. I. Hall, T. A. Green, C. L. Szwarcwald, and N. Pantazis, “Using CD4 Data to Estimate HIV Incidence, Prevalence, and Percent of Undiagnosed Infections in the United States,” JAIDS Journal of Acquired Immune Deficiency Syndromes, vol. 74, no. 1, pp. 3–9, Jan. 2017, doi: 10.1097/QAI.0000000000001151.

[9] I. Hall et al., “HIV Trends in the United States: Diagnoses and Estimated Incidence,” JMIR Public Health and Surveillance, vol. 3, no. 1, p. e8, Feb. 2017, doi: 10.2196/publichealth.7051.

[10] A. Fojo, E. Wallengren, M. Schnure, D. W. Dowdy, M. Shah, and P. Kasaie, “Potential Effects of the Coronavirus Disease 2019 (COVID-19) Pandemic on Human Immunodeficiency Virus (HIV) Transmission: A Modeling Study in 32 US Cities,” Clinical Infectious Diseases, Jan. 2022, doi: 10.1093/cid/ciab1029.

[11] X. Zang et al., “The Potential Epidemiological Impact of Coronavirus Disease 2019 (COVID-19) on the Human Immunodeficiency Virus (HIV) Epidemic and the Cost-effectiveness of Linked, Optout HIV Testing: A Modeling Study in 6 US Cities,” Clinical Infectious Diseases, vol. 72, no. 11, pp. e828–e834, Jun. 2021, doi: 10.1093/cid/ciaa1547.

[12] S. M. Jenness et al., “Projected HIV and Bacterial Sexually Transmitted Infection Incidence Following COVID-19–Related Sexual Distancing and Clinical Service Interruption,” The Journal of Infectious Diseases, vol. 223, no. 6, pp. 1019–1028, Mar. 2021, doi: 10.1093/infdis/jiab051.

[13] A. Satcher Johnson, R. Song, and H. I. Hall, “Estimated HIV Incidence, Prevalence, and Undiagnosed Infections in US States and Washington, DC, 2010–2014,” JAIDS Journal of Acquired Immune Deficiency Syndromes, vol. 76, no. 2, pp. 116–122, Oct. 2017, doi: 10.1097/QAI.0000000000001495.

[14] A. S. Johnson and R. Song, “Incident and Prevalent HIV Infections Attributed to Sexual Transmission in the United States, 2018,” Sexually Transmitted Diseases, vol. 48, no. 4, pp. 285–291, Apr. 2021, doi: 10.1097/OLQ.0000000000001354.

[15] Centers for Disease Control and Prevention, “Estimated HIV incidence and prevalence in the United States, 2010–2015,” HIV Surveillance Supplemental Report 2018 vol. 23(1), 2018. https://www.cdc.gov/hiv/library/reports/hiv-surveillance.html (accessed Jun. 01, 2022).

[16] Centers for Disease Control and Prevention, “Estimated HIV incidence and prevalence in the United States, 2015–2019,” HIV Surveillance Supplemental Report 2021 vol. 26(1), 2021. https://www.cdc.gov/hiv/library/reports/hiv-surveillance.html (accessed Jun. 01, 2022).

[17] Centers for Disease Control and Prevention, “Estimated HIV Incidence in the United States, 2007–2010,” HIV Surveillance Supplemental Report 2012 vol. 17(4), 2012. https://www.cdc.gov/hiv/pdf/library/reports/surveillance/cdc-hiv-surveillance-supplemental-report-vol-17-4.pdf (accessed Jun. 01, 2022).

[18] A. Satcher-Johnson, R. Song, A. Siddigi, A. Hernandez, N. Harris, and D. Daskalasis, “Impact of the COVID-19 Pandemic on HIV Diagnosis in the United States, 2020.”

[19] R. Song and T. Green, “An improved approach to accounting for reporting delay in case surveillance systems,” JP Journal of Biostatistics, vol. 7, no. 1, pp. 1–14, 2012.

[20] N. Crepaz, R. Song, S. B. Lyss, and H. I. Hall, “Estimated time from HIV infection to diagnosis and diagnosis to first viral suppression during 2014–2018,” AIDS, vol. 35, no. 13, pp. 2181–2190, Nov. 2021, doi: 10.1097/QAD.0000000000003008.

[21] Q. Xia et al., “Estimating the probability of diagnosis within 1 year of HIV acquisition,” AIDS, vol. 34, no. 7, pp. 1075–1080, Jun. 2020, doi: 10.1097/QAD.0000000000002510.

[22] J. Skarbinski et al., “Human Immunodeficiency Virus Transmission at Each Step of the Care Continuum in the United States,” JAMA Internal Medicine, vol. 175, no. 4, p. 588, Apr. 2015, doi: 10.1001/jamainternmed.2014.8180.

[23] Z. Li, D. W. Purcell, S. L. Sansom, D. Hayes, and H. I. Hall, “Vital Signs: HIV Transmission Along the Continuum of Care — United States, 2016,” MMWR. Morbidity and Mortality Weekly Report, vol. 68, no. 11, pp. 267–272, Mar. 2019, doi: 10.15585/mmwr.mm6811e1.

[24] HHS Office of Infectious Disease and HIV/AIDS Policy, “What Is Ending the HIV Epidemic in the U.S.?,” Jun. 02, 2021. https://www.hiv.gov/federal-response/ending-the-hiv-epidemic/overview (accessed Aug. 01, 2022).

[25] G. Schwarz, “Estimating the Dimension of a Model,” The Annals of Statistics, vol. 6, no. 2, Mar. 1978, doi: 10.1214/aos/1176344136.

